# Evaluating the Causal Impact of Immune Cells on Breast Cancer and the Mediating Role of Inflammatory Proteins through Mendelian Randomization

**DOI:** 10.1101/2025.01.30.25321182

**Authors:** Qianling Guo, Jiahao Yang, Ying Wen, Kai Zhuang, Aamir Fahira, Lin Yan, Zunnan Huang

**Affiliations:** Key Laboratory of Computer-Aided Drug Design of Dongguan City, The First Dongguan Affiliated Hospital, School of Pharmacy, Guangdong Medical University, Dongguan, P. R. China; Key Laboratory of Big Data Mining and Precision Drug Design of Guangdong Medical University, Key Laboratory for Research and Development of Natural Drugs of Guangdong Province, School of Pharmacy, Guangdong Medical University, Dongguan, P. R. China; School of Biomedical Engineering, Guangdong Medical University, Dongguan, P. R. China; The Second School of Clinical Medicine, Guangdong Medical University, Dongguan, P. R. China

**Keywords:** Breast cancer, Molecular Subtypes, Mendelian Randomization, Immune cells, Mediation analysis, Inflammatory protein

## Abstract

Breast cancer (BC) is a leading cause of mortality among women globally. Emerging evidence suggests that the immune system is involved in BC pathogenesis, with distinct subtypes showing unique immune responses. Inflammation correlates closely with these immune responses. However, the causal role of immune cell characteristics across BC subtypes and the mediating influence of inflammatory proteins remains unclear. We used bi-directional two-sample Mendelian Randomization (MR) to evaluate causal relationships between 731 immune traits and BC risk, including its subtypes, followed by a two-step mediation analysis with 91 inflammatory proteins. The Inverse Variance Weighted (IVW) method served as the primary analysis, supported by sensitivity and reverse MR analyses. Sixteen immune traits showed significant causal associations with BC and its subtypes, with 108 relationships potentially mediated by inflammatory proteins. Traits such as CD25 on CD24+ CD27+, CD25 on IgD- CD38dim, and HLA DR++ monocyte %monocyte acted as protective factors, while CD40 on CD14-CD16+ monocytes posed risks. Notably, CD40L receptor levels mediated up to 73% of the relationship between IgD- CD27-%B cells and ER+BC. Sensitivity analyses indicated no horizontal pleiotropy or reverse causality. This study underscores immune cells’ potential role in BC risk and the mediating impact of inflammatory proteins across subtypes. Targeting specific inflammatory proteins may offer novel strategies for BC risk reduction, informing clinical decision-making and therapeutic development.

## 1. Introduction

Breast cancer (BC) stands as the predominant malignancy afflicting women, presenting a formidable global health dilemma due to its mortality and intricate nature. In the year 2022, the global incidence of BC surpassed 2.30 million cases, resulting in nearly 660,000 deaths [1]. BC is categorized into multiple subtypes based on receptor status and gene expression profiles [2]. Each subtype has a vastly different prognosis and therapeutic response [3], adding to the complexity and uncertainty of treatment [4]. Due to the high heterogeneity of BC, different subtypes of BC are involved in different major functional pathways and immune functions. Therefore, exploring the heterogeneity of immune functions across various BC subtypes can contribute to providing more personalized and precise treatment options for BC patients.

The imbalance of immune cells is intimately linked to the occurrence and development of various cancers, particularly BC. Recent investigations have underscored the association between immune cells and distinct BC subtypes. For instance, a study encompassing 3,771 BC patients undergoing neoadjuvant therapy reported by Denkert [5] highlighted the prognostic significance of Tumor Infiltrating Lymphocytes (TILs) across different subtypes. Sayali et al. [6] discovered significant differences in the composition and function of immune cells among different BC molecular subtypes and observed that notably substantial heterogeneity in prognostic effects between subgroups of ER-positive and ER-negative breast cancer. Moreover, Loi et al. [7] indicate that high levels of TILs are strongly associated with a favorable prognosis in patients with triple-negative breast cancer (TNBC). In contrast, among patients bearing HER2-overexpressing (HER2) lesions, regardless of whether the overexpression is caused by gene amplification, transcriptional or post-transcriptional mechanisms, the prognostic efficacy of TILs is considered suboptimal. Furthermore, Steven et al. demonstrated the efficacy of TILs as a novel immunotherapeutic approach in treating advanced and metastatic breast cancer, confirming the critical role of immunotherapy in breast cancer treatment [8]. Nagarajan et al [9] reveals that a considerable number of immunosuppressive cells, including regulatory T cells (Tregs) and myeloid-derived suppressor cells (MDSC), are present in the tumor microenvironment of BC. These cells play a vital role in hindering effective endogenous immune responses. In some cases, a specific type of immune cell might be detrimental in one context of breast cancer but beneficial in another.

Inflammatory responses play a crucial role in cancer immunity, influencing various stages of tumor development and the effectiveness of cancer therapies. The interaction between inflammation and the immune system can either promote or inhibit tumor growth. For instance, chronic inflammation promotes tumor initiation, progression, and metastasis, and can lead to treatment resistance [10]. Tumor-associated inflammation, primarily driven by innate immune cells, supports tumor growth through the secretion of inflammatory and regenerative cytokines [11]. Furthermore, targeting inflammatory pathways and modulating the immune response are promising strategies for improving cancer therapy outcomes [12]. The intricate crosstalk between immune cells and cancer cells within the tumor microenvironment is crucial for understanding and developing effective cancer therapies. However, the role of immune cells mediated by inflammatory proteins in different molecular subtypes of breast cancer remains largely unexplored. Our study aims to fill this gap by investigating these relationships through Mendelian randomization and mediation analysis.

Mendelian Randomization (MR) is a crucial technique in epidemiological research that provides a powerful analytical tool to uncover genetic predispositions [13]. In this study, we systematically investigate the causal associations between immune traits and overall BC, estrogen receptor status (ER+BC, ER-BC), and molecular subtypes (luminal A-like BC, luminal B-like BC, luminal B/HER2-negative-like BC, HER2-enriched-like BC, TNBC, and BRCA1-mutated TNBC). Furthermore, a two-step, two-sample MR study was used to explore the mediating role of cytokines in the interaction between immune signatures and various types of breast cancer, and multivariate MR (MVMR), Bayesian Weighted MR (BWMR) to assess the causal impact of immune traits on breast cancer risk, to improve the diagnosis and treatment of breast cancer patients.

## 2. Materials and methods

### 2.1 Study design

The study’s flowchart is illustrated in **Figure 1**. The study acquired summary data from Genome-Wide Association Studies (GWAS) for 731 immune traits, along with summary data for breast cancer overall and its eight subtypes. The IVs were rigorously selected before MR analysis. MR must be implemented to satisfy three core assumptions. I) the relevance assumption, which posits a strong association between the IVs and the exposure; II) the independence assumption, indicating that the IVs are not related to any confounders affecting both exposure and outcome; III) the exclusion restriction assumption, meaning that the IVs affect the outcome variable only through the exposure and not via any other pathway [14]. Then, we conducted a bidirectional two-sample MR analysis as well as a series of sensitivity analyses. Furthermore, we performed a two-step MR to screen for mediators of an inflammatory protein in the blood and to quantify each individual mediating effect in the causal associations of the immune traits with breast cancer and its eight subtypes. These comprehensive analyses not only ensure the validity of the causal inferences drawn from the MR approach but also provide a deeper understanding of the underlying mechanisms, particularly by identifying potential mediators and quantifying their effects. This thorough investigation enhances the reliability of the findings and offers new insights into the causal pathways linking immune traits to breast cancer and its subtypes.

**Figure 1.**
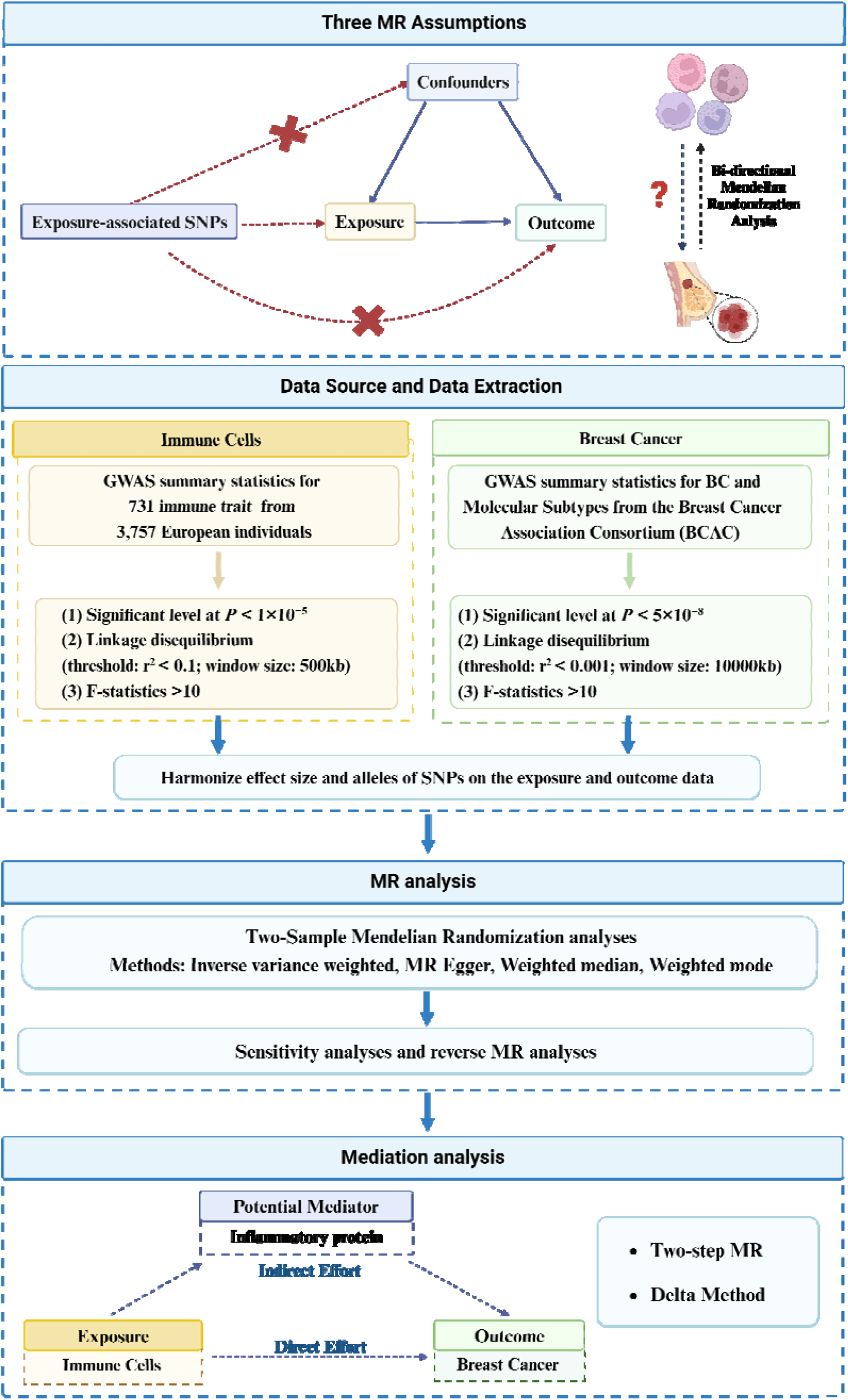
A Schematic diagram of MR Analysis. **Note:** GWAS: Genome-wide association studies; SNP: single-nucleotide polymorphisms (Figure created with biorender.com).

### 2.2 Data sources for immune traits

To obtain a more comprehensive and reliable conclusion regarding the causal association between immune indicators and BC, this study utilized the largest available GWAS data for immunophenotyping of peripheral blood. The study covers a total of 731 immune phenotypes, including Absolute Cell (AC) counts, Median Fluorescence Intensities (MFI) measurements reflecting surface antigen levels, Morphological Parameters (MP, forward scatter (FSC), and side scatter (SSC)), which are proportional to cell volume, intracellular complexity, and cell surface texture, and Relative Cell (RC) counts. These immune phenotypes can be categorized by the panel into seven groups (B cell, TBNK, Monocyte, cDC, Maturation stages of T cell, Treg, Myeloid cell). Statistical data is publicly available in the GWAS catalog [15]. The GWAS analysis is based on 3,757 Sardinian samples (57% female), testing approximately 22 million single nucleotide polymorphisms (SNPs) genotyped through high-density arrays after adjusting for sex, age, and the square of age [15]. These SNPs were imputed using a Sardinian sequence-based reference panel [16].

#### Data sources for breast cancer

In order to gain a comprehensive understanding of the causal association between BC risk and immune indicators, summary statistics of genetic associations from the Breast Cancer Association Consortium (BCAC) (consisting of 68 studies combined together) [17], as well as the Discovery, Biology and Risk of Inherited Variants in Breast Cancer Consortium (DRIVE), were collected. Concisely, these data encompass 122,977 BC cases and 105,974 controls, further stratified by estrogen receptor (ER) expression into ER-positive BC cases and ER-negative BC cases (**Table 1**). Genotyping was conducted using the iCOGS array or the OncoArray, and imputation was performed using the third phase of the 1000 Genomes Project dataset, yielding data on 10,680,257 SNPs. The results were then summarized using a fixed-effect meta-analysis [17].

**Table 1.** GWAS data sources for breast cancer.

| Breast cancer subtype | Cases/Controls | Ancestry | Source (PMID) |
| --- | --- | --- | --- |
| Overall BC risk | 122,977/105,974 | European | 29059683 |
| ER+BC risk | 69,501/105,974 | European | 29059683 |
| ER-BC risk | 21,468/105,974 | European | 29059683 |
| Luminal A BC risk | 7325/20,815 | European | 32424353 |
| Luminal B BC risk | 1682/20,815 | European | 32424353 |
| Luminal B and HER2 negative BC risk | 1779/20,815 | European | 32424353 |
| HER2-enriched BC risk | 718/20,815 | European | 32424353 |
| Triple-negative BC risk | 2006/20,815 | European | 32424353 |
| BRCA1 mutated triple negative | 18,016/100,971 | European | 32424353 |

Additionally, to delve deeper into the impact of immune indicators on the risk associated with different molecular subtypes of BC, summary statistics were derived from a dataset [18] comprising 118,474 cases and 96,201 controls. These data were previously examined across 82 studies by BCAC, aiming to obtain associations’ summary statistics for five BC subtypes (**Table 1**). Concisely, these subtypes are based on the expression of ER, progesterone receptor (PR), human epidermal growth factor receptor 2 (HER2), and cancer grade: luminal A-like (ER+ and/or PR+, HER2- and grade 1/2), luminal B-like (ER+ and/or PR+, HER2+), luminal B/HER2-negative-like (ER+ and/or PR+, HER2-, grade 3), HER2-enriched-like (ER-, PR-, HER2+), and TNBC (ER-, PR-, HER2-). Additionally, summary data [18] from the Consortium of Investigators of Modifiers of BRCA1/2 (CIMBA), comprising cases with BRCA1 mutation and corresponding controls without BRCA1 mutation, were included in the analysis. As the majority of BRCA1-mutated cancers were also triple-negative, we utilized data from meta-analyses on cancers with BRCA1 mutations and TNBC, based on summary statistics [18].

### Data sources for Inflammatory protein

Inflammatory protein data were obtained from a study by Zhao et al. [19], which conducted a genome-wide protein quantitative trait loci (pQTL) analysis of 91 inflammation-related plasma proteins. This analysis included 11 cohorts, encompassing a total of 14,824 participants of European ancestry. The dataset provides a comprehensive genetic mapping of proteins implicated in inflammatory processes, offering valuable insights into their regulatory mechanisms. This dataset was utilized for mediation analyses to investigate the relationships between immune cells, inflammatory proteins, and breast cancer, including its various subtypes. By leveraging this data, we aimed to elucidate the potential pathways linking immune traits to breast cancer outcomes.

### 2.3 Selection of instrumental variables

To thoroughly investigate the relationship between breast cancer (BC) and its subtypes with immune traits, the significance threshold was widened to *P* < 1×10^−5^ [20]. Furthermore, for BC and its subtypes, we utilized SNPs that met the genome-wide significance threshold (*P* < 5×10^−8^) as genetic instruments. To ensure the independence of SNP sites, linkage disequilibrium (LD) analysis is performed using the R package ‘TwoSampleMR’. The filtering criteria for immune traits are set at r^2^=0. 1 and kb=500, indicating the exclusion of SNPs within a 500 kb range that has an r^2^ greater than 0.1 with the most significant SNP. For the overall BC and its molecular subtypes and inflammatory protein, the filtering criteria are r^2^=0.001 and kb=10000 [21, 22]. The R package ‘TwoSampleMR’ is used for consistency analysis of the effect alleles of SNPs related to the exposure factor and outcome phenotype, ensuring the consistency of the effect alleles and removing all SNPs with palindromic structures. Furthermore, to assess the strength of the selected SNPs, the following formula is used to calculate the corresponding R^2^ and F-statistic for each SNP, and SNPs with an F-statistic < 10 are discarded to avoid introducing weak instrument variable bias [21, 23] (**Supplementary Table S1**-**S3**).

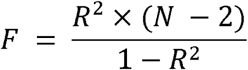

R^2^ indicates the IV explains the degree of exposure, also known as PVE (phenotypic variance explained), and N represents the sample size.

### 2.4 MR analyses

The primary analysis of the study involves using the inverse-variance weighted method (IVW) [24] to examine the relationship between each immune trait and both overall breast cancer (BC) and its subtypes. Subsequently, the odds ratio (OR) and 95% confidence intervals (CI) are calculated for these associations. Concisely, the effect sizes and standard errors for exposure and outcome are obtained for each genetic variant. Then, a weighted sum of the effects represented by the genetic instruments on the outcome is calculated to determine the overall effect size. In addition, this study conducts multiple tests, such as weighted median [25], weighted mode [26], and MR-Egger [27]. Furthermore, a bi-directional MR analysis was conducted to investigate the presence of reverse causal relationships.

### 2.5 Multivariable Mendelian Randomization

Multivariable Mendelian Randomization (MVMR) analysis provides a robust framework for estimating the direct causal effects of one or more exposures. It serves as a powerful tool for establishing causal relationships across diverse scenarios, utilizing summary-level data [28]. In MVMR, the instruments may influence multiple risk factors, as long as they meet the necessary instrumental variable assumptions [29]. In this study, we applied the MVMR, incorporating all available instrumental variables related to immune cells, to assess their independent effects on BC. Analyses were conducted using Inverse Variance Weighted (IVW) and MR-Egger regression methods, with the MR-Egger intercept used to identify potential horizontal pleiotropy.

### 2.6 Bayesian Weighted Mendelian Randomization

Bayesian Weighted Mendelian Randomization (BWMR) integrates Bayesian statistics with MR to account for the uncertainty associated with weak effects estimates in GWAS and the potential influence of horizontal pleiotropy. This approach enhances the robustness of the analysis by providing both computational stability and statistical efficiency. In our study, we utilized BWMR to validate the findings obtained through the Inverse Variance Weighted (IVW) method [30].

### 2.7 Mediation MR analysis

A two-step MR analysis was performed to investigate the potential mediating role of inflammatory proteins in the association between the immune cell and breast cancer. In the first step, univariable MR (UVMR) was employed to determine the causal effect of the genetically determined immune cell and inflammatory proteins (β1). The second step involved estimating the causal impact of each inflammatory protein as a mediator on breast cancer and its subtypes (β2), based on the assumption that the mediator is causally linked to the UVMR outcome. The mediation proportion of each mediator in the association between the immune cell and breast cancer was calculated as:

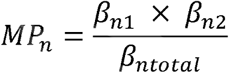

where β_n1_ represents the causal effect for each immune cell-inflammatory protein pair, β_n2_ represents the causal effect for each inflammatory protein/breast cancer pair, β_ntotal_ represents the total causal effect for each immune cells/breast cancer pair, and *MPn* represents the mediation proportion for each pair [31]. Confidence intervals were estimated using the delta method [32].

### 2.8 MR sensitivity analysis

This study conducted a series of sensitivity analyses to assess the robustness of the results. Concisely, Heterogeneity was evaluated using Cochran’s Q test, with a *P-value* < 0.05 indicating the presence of heterogeneity [33]. The MR-Egger intercept test was utilized to detect the influence of horizontal pleiotropy on causal association estimates, with a *P-value* < 0.05 indicating the presence of horizontal pleiotropy [34]. The reliability of the association was assessed through leave-one-out analysis, funnel plots, and scatter plots [35]. Briefly, a leave-one-out analysis was performed to ensure that the findings were not influenced by any individual SNP, while scatter plots were used to show that the results were not driven by outliers. Funnel plots are utilized to evaluate the reliability of the association. Furthermore, to account for the possibility of false positives, the Benjamini-Hochberg False Discovery Rate (FDR) correction was applied [36]. We considered unadjusted IVW method results with *P-value* < 0.05 as indicative of an association, while FDR-adjusted IVW method results with *P_FDR_* < 0.05 were considered as a significant association between exposure and outcome. Furthermore, an association was considered suggestive when *P_FDR_*< 0.10.

### 2.9 Statistical analysis

All analyses were performed in R software version 4.3.1 (http://www.Rproject.org). IVW, weighted median, and MR-Egger were conducted using the “TwoSampleMR” package (version 0.5.7).

## 3. Results

### 3.1 Overview of Mendelian randomization

In this study, MR analysis was performed on 731 immune cell phenotypes of BC and its molecular subtypes, with a total of 6579 associations (731 exposures×9 outcomes) included (**Supplementary Table S4**), and a total of 345 pairs reached nominal significance in the IVW analysis (*P-value*<0.05). Given that immune traits were divided into four types of phenotypes (MFI, AC, RC, and MP) including seven panels of immune cells, the detected potential immune signatures associated with BC can be summarized into 340 MFIs, 88 ACs, 143 RCs, and 19 MP traits (with some overlapping signals shared by all types of BC at the four sites). When the immunophenotypes were categorized according to Panel, 57 traits belonged to the maturation stages of T cell, 98 to TBNK, 115 to Treg, 148 to B cell, 44 to Myeloid cell, 71 to cDC, and 57 to Monocyte were found to be suggestively associated with BC.

After multiple test corrections and sensitivity analyses, 16 pairs of associations, including 13 MFIs (a phenotype shared by Luminal A BC and BRCA1 mutated TNBC), 1 AC, and 1 RC, remained statistically significant (**Figure 2 and Table 3**), and 18 pairs suggested a suggestive association between exposure and outcome (**Supplementary Figure S1**).

**Figure 2.**
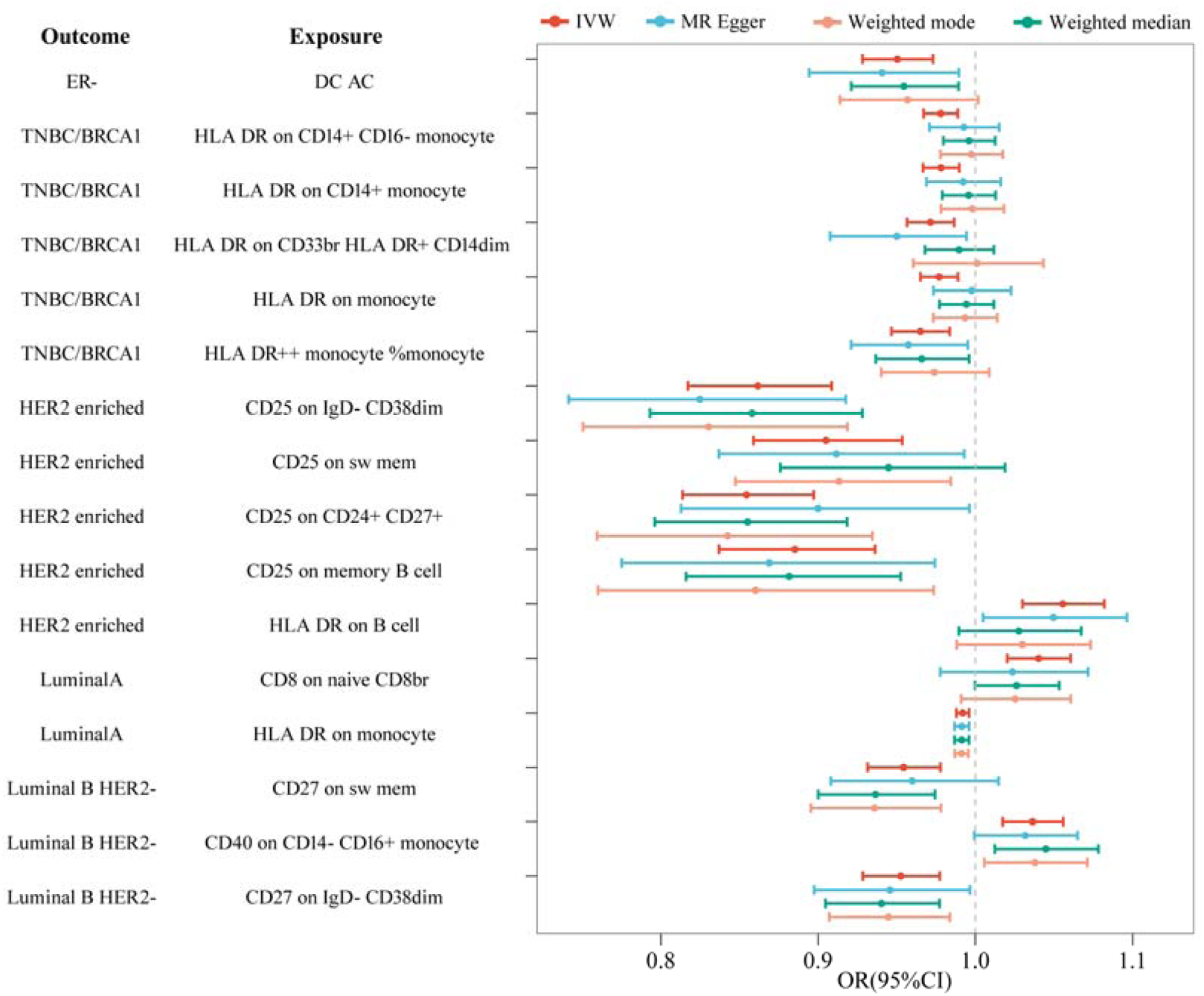
Forest plot revealing the significant association between immune traits and breast cancer and its molecular subtypes through different approaches. **Note:** CI: Confidence Interval; OR: Odds Ratio; ER-: ER-BC; Luminal A: Luminal A like BC; Luminal B HER2-: Luminal B/HER2-negative-like BC; HER2 enriched: HER2-enriched-like BC; TNBC/BCRA1: BRCA1-mutated TNBC.

### 3.2 Two-sample MR analysis of immune traits and breast cancer risk

By performing MR analysis on the overall BC, the results showed a total of 51 pairs of indicative associations (*P-value* < 0.05) in the overall BC according to the results of the IVW method (**Supplementary Table S4**). However, no immune traits were identified as significant and suggestive associations after correction using the FDR method.

### 3.3 Two-sample MR analysis of immune traits and ER-defined typing of breast cancer

An immunophenotype was identified in ER-BC (Estrogen-negative breast cancer) by the FDR method. Results from the IVW method indicated that DC AC significantly decreased the risk of ER-BC (cDC; OR: 0. 95, 95% CI: 0. 93-0. 97, *P-value*= 2. 38×10^-5^, *P_FDR_* = 0. 017), which was also confirmed by the MR-Egger (OR: 0. 94, 95% CI: 0. 89-0. 99, *P-value*= 0. 021) and Weighted median methods (OR: 0. 95, 95%CI: 0. 92-0. 99, *P-value*= 0. 010) (**Figure 2 and Table 2**). In addition, no immunophenotypes were detected for ER+BC (Estrogen-positive breast cancer) after the FDR approach. Only 1 immune cell phenotype was identified as potentially linked to ER+BC, presenting a suggestive association with a significance level of 0. 064. CD20 on switched memory (sw mem) (OR: 1.03, 95CI:1.01-1.04; *P-value*= 8. 79×10^-5^; B cell) (**Supplementary Figure S1** and **Supplementary Table S4**).

**Table 2.** Significant associations of immune cell traits with the risk of ER-defined typing of breast cancer: Findings from Mendelian randomization analyses.

| Outcome | Exposure | nSNP | Methods | OR (95%CI) | $P\text{-value}$ | $P_{FDR}$ |
| --- | --- | --- | --- | --- | --- | --- |
| ER- | DC AC | 51 | IVW | 0.95(0.93-0.97) | $2.38 \times 10^{-5}$ | 0.017 |
|  |  | 51 | MR Egger | 0.94(0.89-0.99) | 0.021 |  |
|  |  | 51 | Weighted median | 0.95(0.92-0.99) | 0.009 |  |
|  |  | 51 | Weighted mode | 0.96(0.91-1.00) | 0.078 |  |

**Note:** nSNP: Number of single-nucleotide polymorphisms; CI: Confidence Interval; OR: Odds Ratio; IVW: inverse-variance weighted. ER-: ER-BC

**Table 3.**
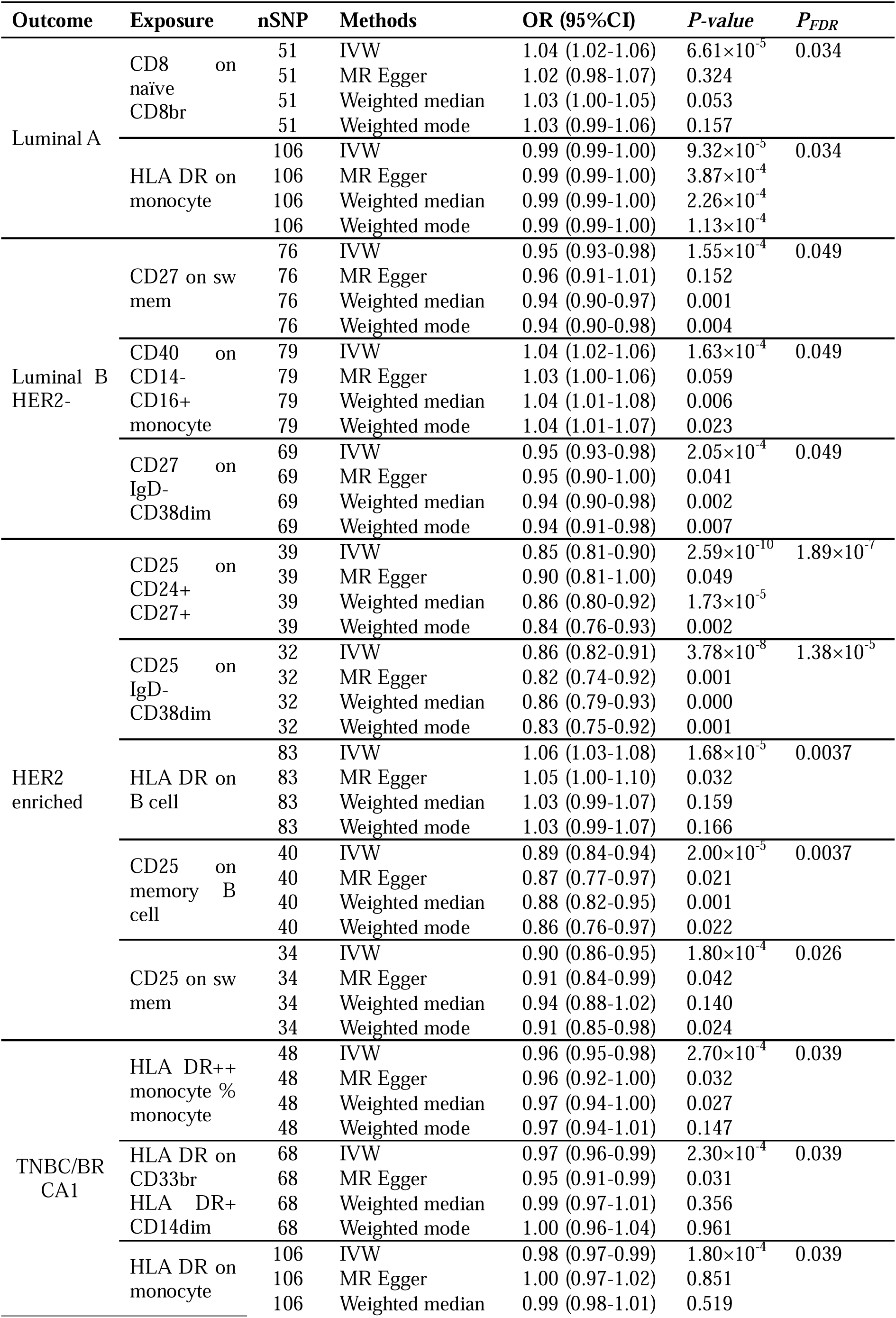
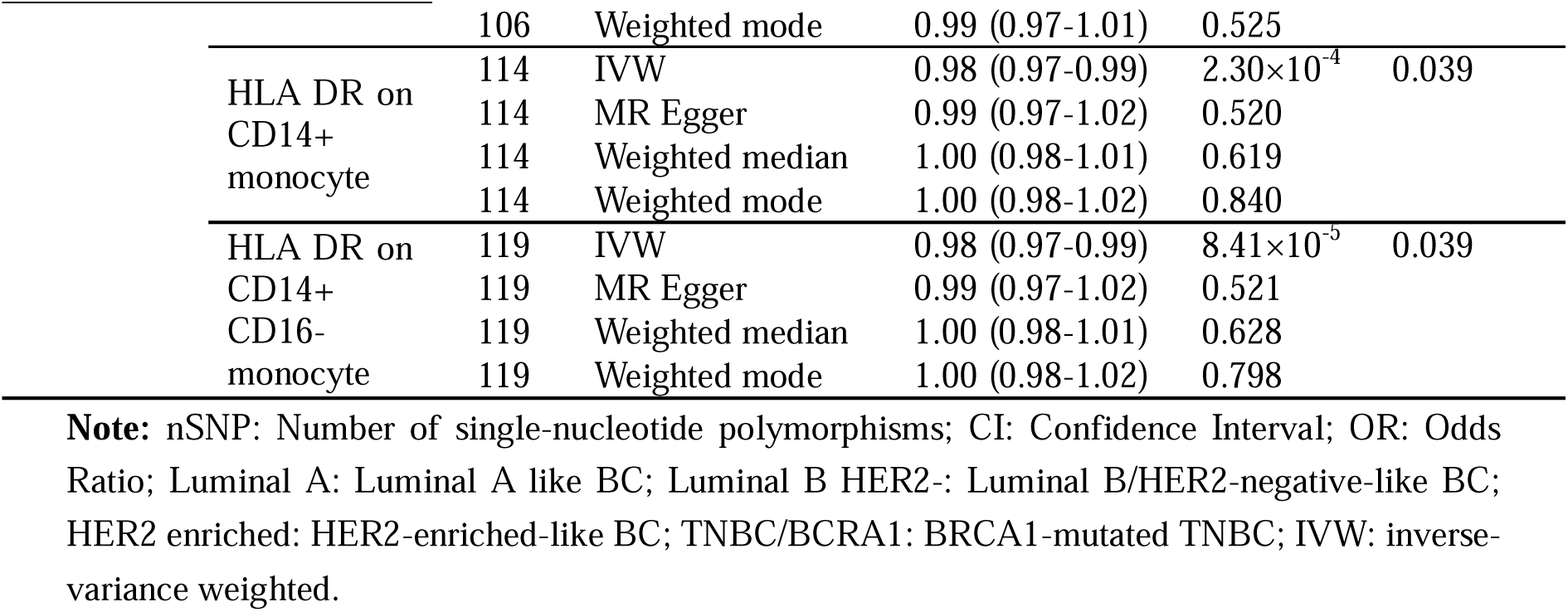
Significant associations of immune cell traits with the risk of molecular subtypes of breast cancer: Findings from Mendelian randomization analyses.

### 3.4 Two-sample MR analysis of immune traits and molecular subtypes of breast cancer

In this study, Luminal A-like BC was significantly associated with two immunophenotypes by the IVW method after the adjustment of the FDR method (**Figure 2** and **Table 3**). Results from the IVW method indicated that CD8 on naive CD8br (Maturation stages of T cell; OR: 1. 04, 95%CI: 1. 02-1. 06, *P-value=* 6. 61×10^-5^, *P_FDR_* = 0. 034) and HLA DR on monocyte (Monocyte; OR: 0. 99, 95%CI: 0. 99-1. 00, *P-value**=*** 9. 32×10^-5^, *P_FDR_* = 0. 034) significantly increased the risk of Luminal A BC, and other MR methods also confirmed a positive association (**Figure 2** and **Table 3**). Meanwhile, this study detected suggestive associations between seven immune traits and Luminal A-like BC (*P_FDR_* < 0. 1). Notably, CD20 on sw mem (B cell; *P-value*= 0.001, OR:1.03) (**Supplementary Figure S1** and **Supplementary Table S4**) showed the same results in ER+BC, both as potential risk factors. Moreover, CD27 on sw mem, HLA DR on CD14+ CD16-monocyte, and HLA DR on CD14+ monocyte (**Supplementary Figure S1** and **Supplementary Table S4**), which present a potential protective effect against Luminal A-like BC, were also significant in other molecular typing of BC.

In this study, three immunophenotypes of Luminal B/HER2-negative-like BC were identified after the FDR method. Concisely, CD27 on sw mem (OR: 0. 95, 95%CI: 0. 93-0. 98, *P-value*= 1. 55×10^-4^), and CD27 on IgD- CD38dim (OR: 0. 95, 95CI%: 0. 93-0. 98, *P-value=* 2. 1×10^-4^) was associated with a decreased risk of Luminal B/HER2-negative-like BC, both phenotypes belonging to the B cell group, with a *P_FDR_*of 0. 0499, while other MR methods similarly supported a negative correlation of the results (**Figure 2** and **Table 3**). In addition, there was a positive correlation between CD40 on CD14-CD16+ monocyte and Luminal B/HER2-negative-like BC (Monocyte; OR: 1. 04, 95%CI: 1. 02-1. 06, *P-value=* 1. 6×10^-4^, *P_FDR_* = 0. 0499) (**Figure 2** and **Table 3**).

After FDR correction, the IVW MR analyses identified five immunophenotypes detected to be significantly associated with HER2-enriched-like BC (**Figure 2** and **Table 3**). The B cell group exhibited four predominant phenotypes, all linked to a reduced risk of HER2-enriched-like breast cancer. (CD25 on CD24+ CD27+, OR: 0.85, 95%CI: 0.81-0.90; CD25 on IgD- CD38dim, OR: 0.86, 95%CI: 0.82-0.91; CD25 on memory B cell, OR: 0.89, 95%CI: 0.84-0.94; CD25 on sw mem, OR: 0.90, 95%CI:0.86-0.95) (**Figure 2** and **Table 3**). In contrast, HLA DR on B cell was associated with an elevated risk of HER2-enriched-like BC (TBNK; OR: 1. 06, 95%CI: 1. 03-1. 08, *P-value=* 1. 68×10^-5^, *P_FDR_* = 0. 0037), and the results were similarly supported by other MR methods (**Figure 2** and **Table 3**). Meanwhile, a suggestive immunophenotype, Granulocyte AC located on TNBK (OR: 0.88, 95%CI: 0.82-0.95, *P-value*= 4.8×10^-4^, *P_FDR_* = 0.0587), was also detected to be negatively correlated with HER2-enriched-like BC, which was found to be in the same direction of correlation with the results using other MR methods (**Supplementary Figure S1** and **Supplementary Table S4)**.

As for BRCA1-mutated TNBC, this study found that five immunophenotypes were protective after FDR correction. The IVW method results suggested a negative correlation between HLA DR expression on CD14+ CD16-monocytes, HLA DR expression on monocytes, HLA DR expression on CD14+ monocytes, and BRCA1-mutated TNBC, with *P_FDR_* of 0.039 respectively (**Figure 2** and **Table 3**). Furthermore, HLA DR on CD33br HLA DR+ CD14dim and HLA DR++ monocyte %monocyte were associated with a decreased risk of BRCA1-mutated TNBC (**Figure 2** and **Table 3**). All significant phenotypes were observed to be HLA DR, indicating the critical role of these phenotypes in BRCA1-mutated TNBC. Furthermore, this study also found that HLA DR on monocytes was significantly associated with both Luminal A-like BC and BRCA1-mutated TNBC, which is important for revealing the disease mechanism and potential common therapeutic targets. In addition, nine immunophenotypes showed suggestive associations with BRCA1-mutated TNBC, among which the increase of DC AC in cDC was protective against BRCA1-mutated TNBC (**Supplementary Figure S1** and **Supplementary Table S4**), which showed the same inverse association in ER-BC. Notably, CD25 on IgD+ and CD24 on transitional in the B cell group were potential risk factors for BRCA1-mutated TNBC (**Supplementary Figure S1** and **Supplementary Table S4**).

Moreover, the results obtained through the IVW method, following FDR correction, did not reveal any significant or suggestive association between immune cell phenotypes and Luminal B-like BC and TNBC.

### 3.5 BWMR analysis between immune traits on breast cancer

Given the diversity of the study population and the complexity of the data, we used BWMR analyses to more effectively control for potential confounders and to improve the accuracy and robustness of causal inferences. The results showed that all 16 immunophenotypes were significantly causally associated with BC. Detailed information can be found in **Supplementary Table S5**.

### 3.6 MVMR analysis between immune traits on breast cancer

To determine whether significant immunophenotypes have an effect on breast cancer risk directly or by mutual adjustment, we further performed an MVMR analysis. The effect of HLA DR++ monocyte %monocyte on BRCA1-mutated TNBC remained after accounting for reciprocal adjustments for significant immunophenotypes. The association between Luminal B/HER2-negative-like BC, Luminal A-like BC, and HER2-enriched-like BC was then attenuated after significant immunophenotypic cross-adjustment. Detailed information can be found in **Supplementary Table S6**.

### 3.7 Sensitivity analysis

Horizontal pleiotropy between the 16 pairs of immune cell phenotypes screened above and BC was examined by MR Egger intercept, and no pleiotropy of immune phenotypes was detected (**Supplementary Table S7**). Heterogeneity was subsequently assessed using Cochran’s Q test, revealing that 11 of these immunophenotype pairs did not exhibit heterogeneity (**Supplementary Table S7**), namely CD25 on CD24+ CD27+, CD25 on IgD- CD38dim, HLA DR on B cell, DC AC, CD25 on sw mem, HLA DR on CD14+ CD16-monocyte, HLA DR on CD33br HLA DR+ CD14dim, HLA DR++ monocyte %monocyte, CD27 on sw mem, CD40 on CD14-CD16+ monocyte, CD27 on IgD- CD38dim. Leave-one-out sensitivity analysis for each pair of associations is shown in **Figure S2-S6**, while scatterplots and funnel plots for each pair of associations are shown in **Figure 3** and **Figure 4**.

**Figure 3.**
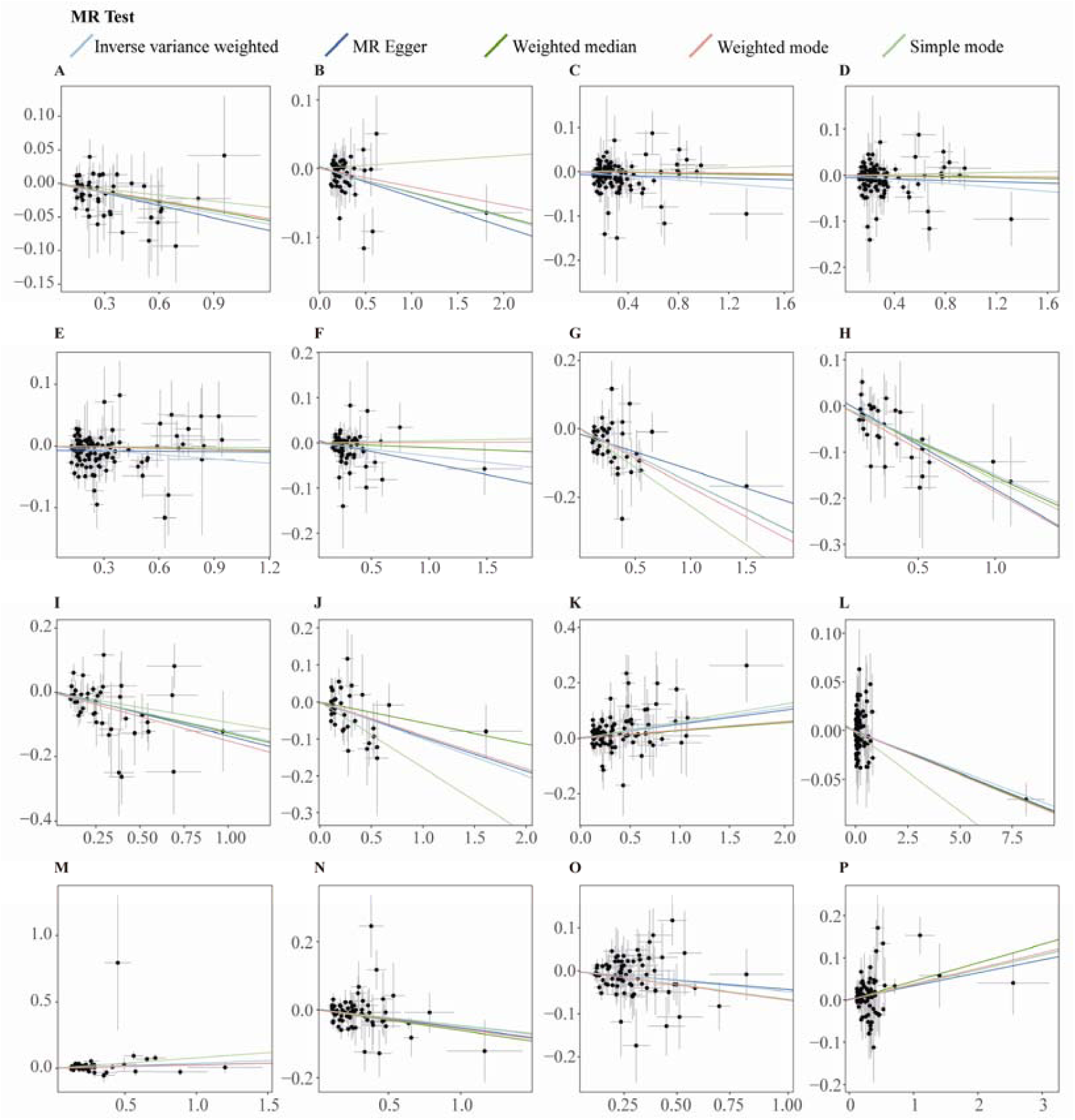
Scatter plot of a two sample Mendelian randomization study of immune traits and BC. **(A)** DC AC on ER-BC. **(B)** HLA DR++ monocyte %monocyte on BRCA1-mutated TNBC. **(C)** HLA DR on CD14+ CD16-monocyte on BRCA1-mutated TNBC. **(D)** HLA DR on CD14+ monocyte on BRCA1-mutated TNBC. **(E)** HLA DR on monocyte on BRCA1-mutated TNBC. **(F)** HLA DR on CD33br HLA DR+ CD14dim on BRCA1-mutated TNBC. **(G)** CD25 on IgD- CD38dim on HER2-enriched-like BC. **(H)** CD25 on IgD- CD38dim on HER2-enriched-like BC. (I) CD25 on CD24+ CD27+ on HER2-enriched-like BC. **(J)** CD25 on sw mem on HER2-enriched-like BC. (K) HLA DR on B cell on HER2-enriched-like BC. **(L)** HLA DR on monocyte on Luminal A-like BC. (M) CD8 on naive CD8br on Luminal A-like BC. **(N)** CD27 on IgD- CD38dim on Luminal B/HER2-negative-like BC. **(O)** CD27 on sw mem on Luminal B/HER2-negative-like BC. **(P)** CD40 on CD14-CD16+ monocyte on Luminal B/HER2-negative-like BC.

**Figure 4.**
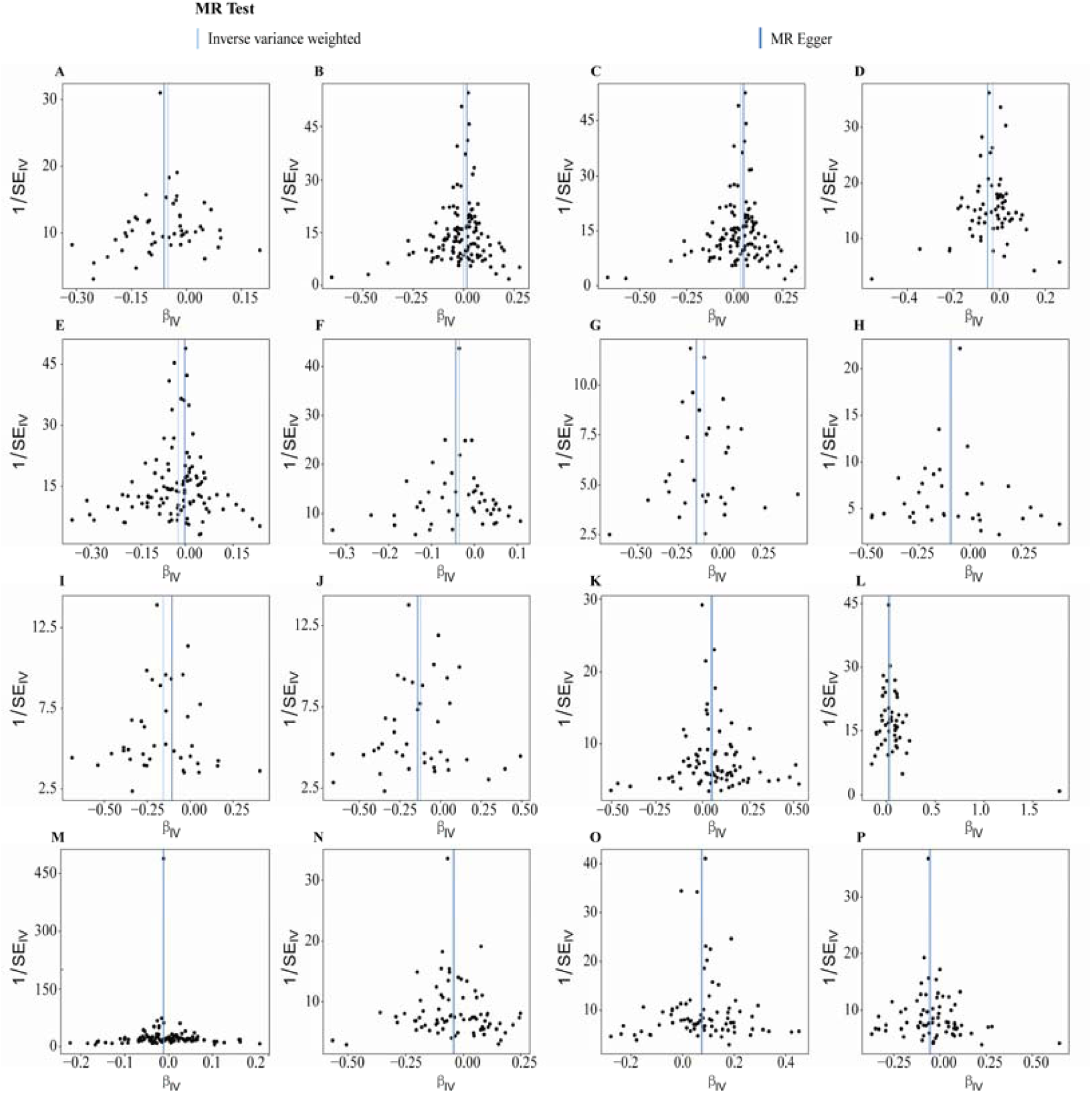
Funnel plot of a two-sample Mendelian randomization study of immune traits and BC. **(A)** DC AC on ER-BC. **(B)** HLA DR on CD14+ CD16-monocyte on BRCA1-mutated TNBC. **(C)** HLA DR on CD14+ monocyte on BRCA1-mutated TNBC. **(D)** HLA DR on CD33br HLA DR+ CD14dim on BRCA1-mutated TNBC. **(E)** HLA DR on monocyte on BRCA1-mutated TNBC. (F) HLA DR++ monocyte %monocyte on BRCA1-mutated TNBC. **(G)** CD25 on IgD- CD38dim on HER2-enriched-like BC. **(H)** CD25 on sw mem on HER2-enriched-like BC. **(I)** CD25 on CD24+ CD27+ on HER2-enriched-like BC. **(J)** CD25 on memory B cell on HER2-enriched-like BC. (K) HLA DR on B cell on HER2-enriched-like BC. **(L)** CD8 on naive CD8br on Luminal A-like BC. **(M)** HLA DR on monocyte on Luminal A-like BC. **(N)** CD27 on sw mem on Luminal B/HER2-negative-like BC. **(O)** CD40 on CD14-CD16+ monocyte on Luminal B/HER2-negative-like BC. **(P)** CD27 on IgD- CD38dim on Luminal B/HER2-negative-like BC.

### 3.7 Reverse MR analysis of breast cancer on immune traits

Based on inverse MR analysis and FDR correction, only a significant association was found between TNBC and CD3 on TD CD4+ (Maturation stages of T cell; IVW OR: 0. 74, 95%CI: 0. 65-0. 84, *P-value*= 3. 82×10^-6^, *P_FDR_* = 0. 0028) (**Figure 5** and **Supplementary Table S8**). No other significant association was found between BC and molecular subtypes and immunophenotypes. Sensitivity analysis of the results for significance, Cochran’s Q test did not reveal heterogeneity and MR Egger’s intercept test did not detect horizontal pleiotropy. (**Supplementary Table S9**)

**Figure 5.**
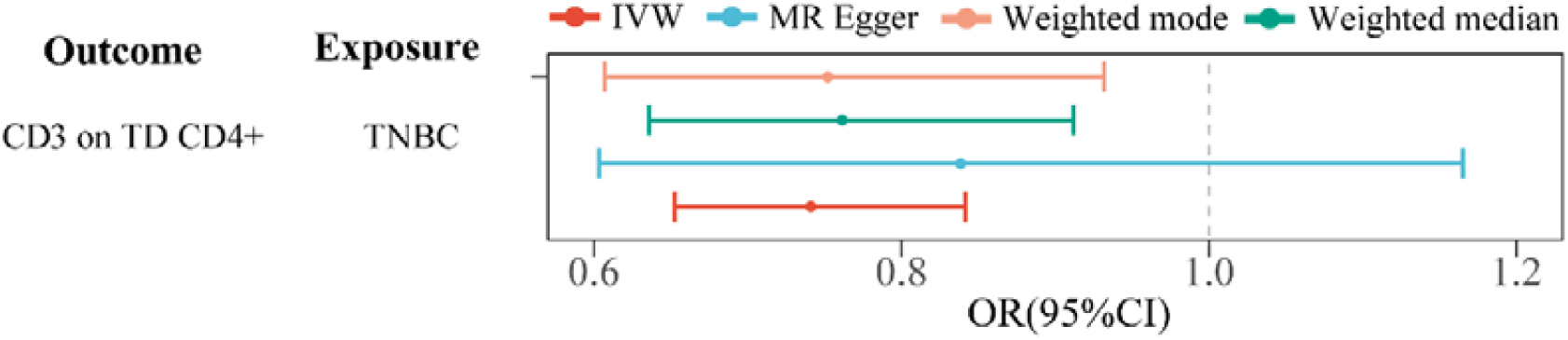
Forest plot revealing the significant association between breast cancer and its molecular subtypes and immune traits through different approaches. **Note:** CI: confidence interval; OR: Odds Ratio; TNBC: Triple-negative breast cancer

### 3.8 Effect of inflammatory protein on breast cancer

By performing MR analysis between inflammatory proteins and BC, 66 significant associations were identified. A key finding revealed that higher levels of the leukemia inhibitory factor receptor were associated with a decreased risk of developing Luminal A-like BC (P-value = 7.9 × 10CC, OR: 0.89, 95% CI: 0.84–0.95) (**Supplementary Figure S7**). All MR results are presented in **Supplementary Table S10**.

### 3.9 Mediation analysis of immune traits on breast cancer

By conducting a mediation MR analysis, we investigated whether inflammatory proteins mediate the impact of immunophenotype on BC. For the causal association between immune traits and Overall BC, ER+BC, and Luminal A-like BC, the highest mediation proportion was predicted for CD40L receptor levels, accounting for 64%, 73% and 68% (**Table 4**) respectively. In addition, Interleukin-15 receptor subunit alpha in HLA DR on CD14+ CD16-monocyte and Luminal B/HER2-negative-like BC showed the highest mediation proportion of 39% (**Table 4**). Furthermore, In the causal relationship between CD3 on CM CD4+ T cells and ER-BC, matrix metalloproteinase-1 levels accounted for the highest mediation proportion, reaching 17% (**Table 4**). For the causal relationship between CD39+ resting T regulatory cells and TNBC, TNF-related activation-induced cytokine levels accounted for the highest mediation proportion at 10% (**Table 4**). Similarly, in the causal relationship between CD64 on CD14+ CD16+ monocytes and Luminal B like BC, fibroblast growth factor 19 levels accounted for the highest mediation proportion, reaching 12% (**Table 4**). For the causal association between BAFF-R on IgD+ CD38dim B cells and HER2-enriched-like BC, C-X-C motif chemokine 10 levels represented the highest mediation proportion, accounting for 20% (**Table 4**). Detailed information can be found in **Supplementary Table S11** and **Supplementary Figure S8-S12.**

**Table 4.**
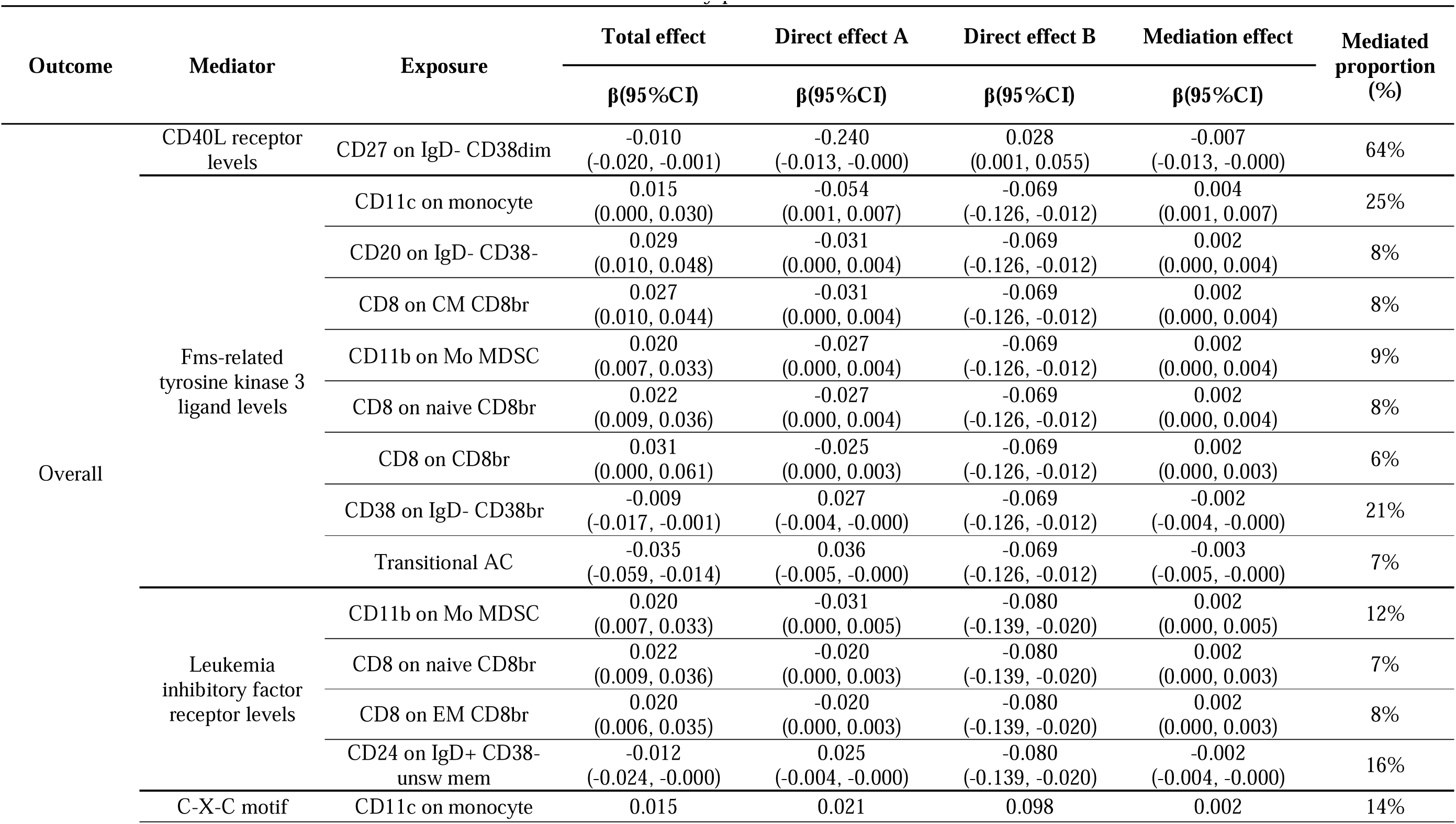
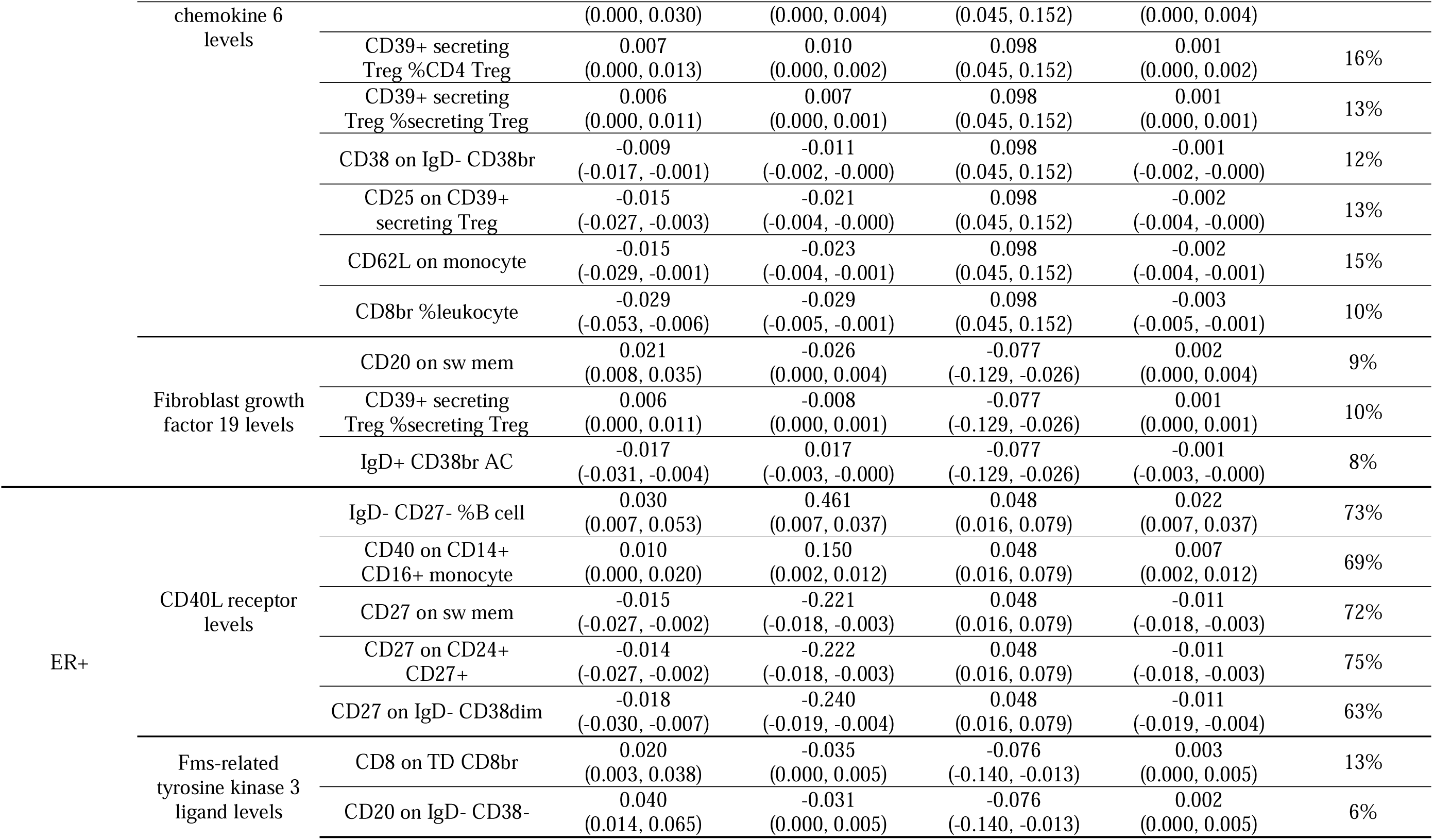
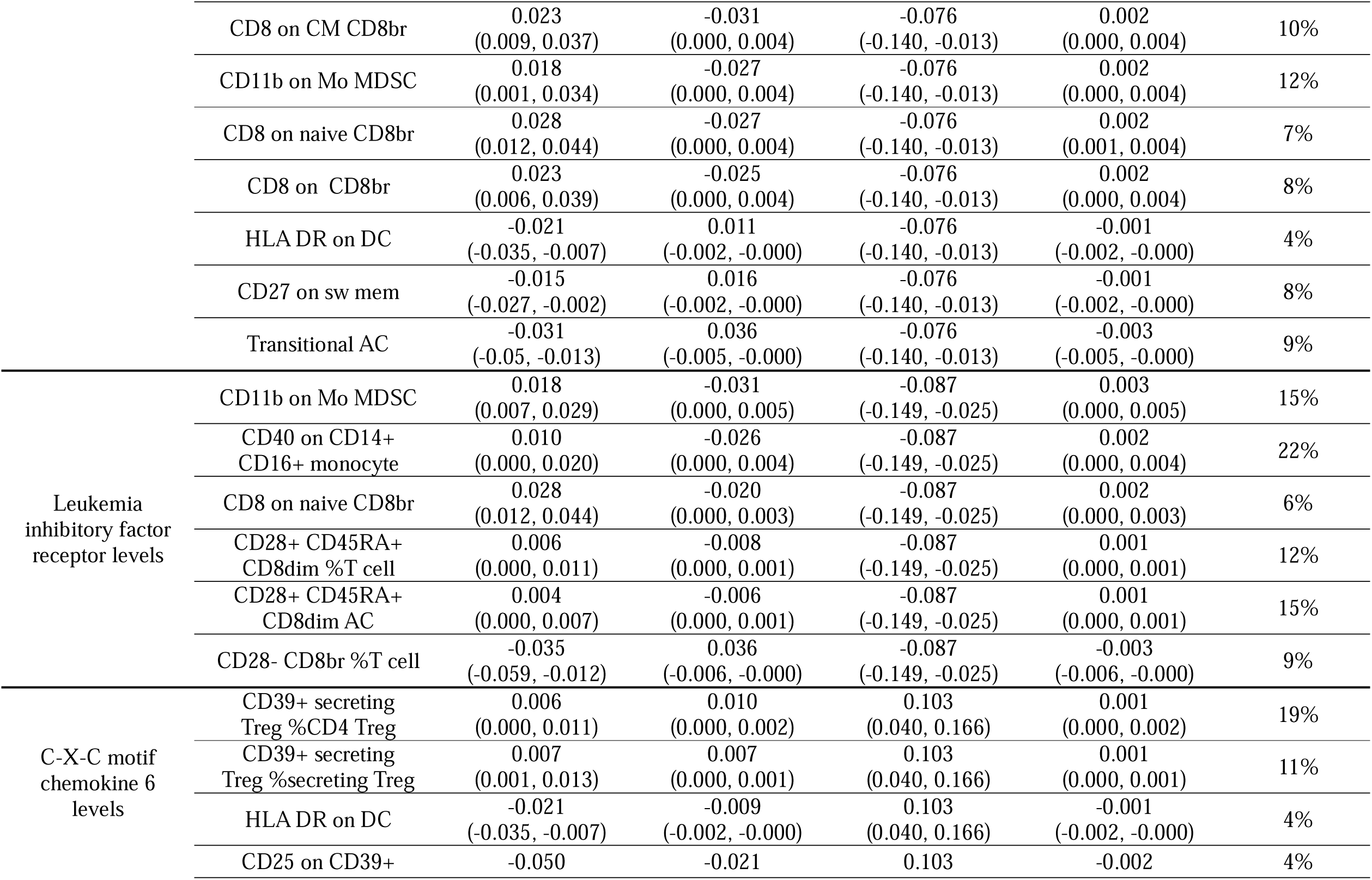
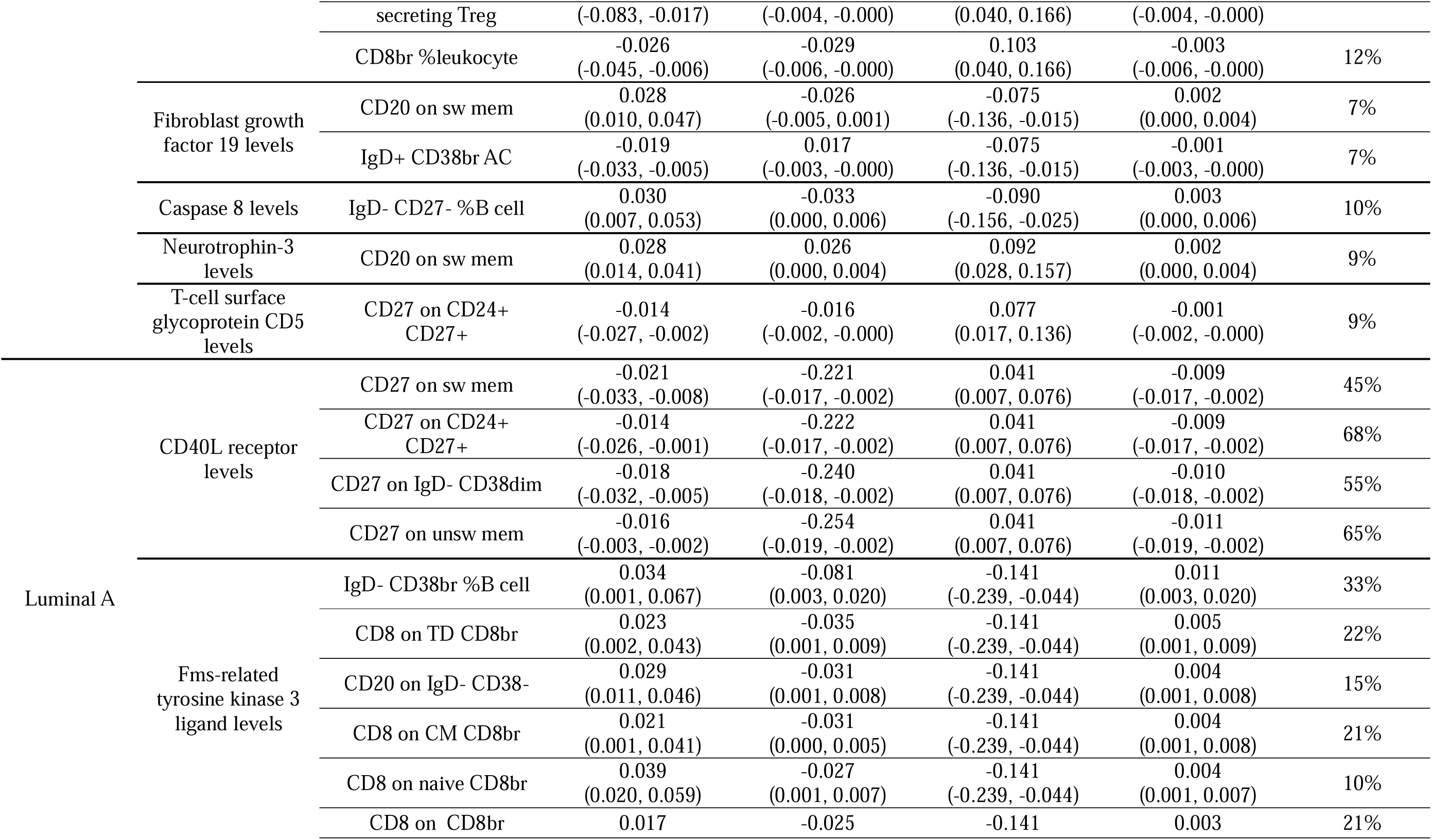
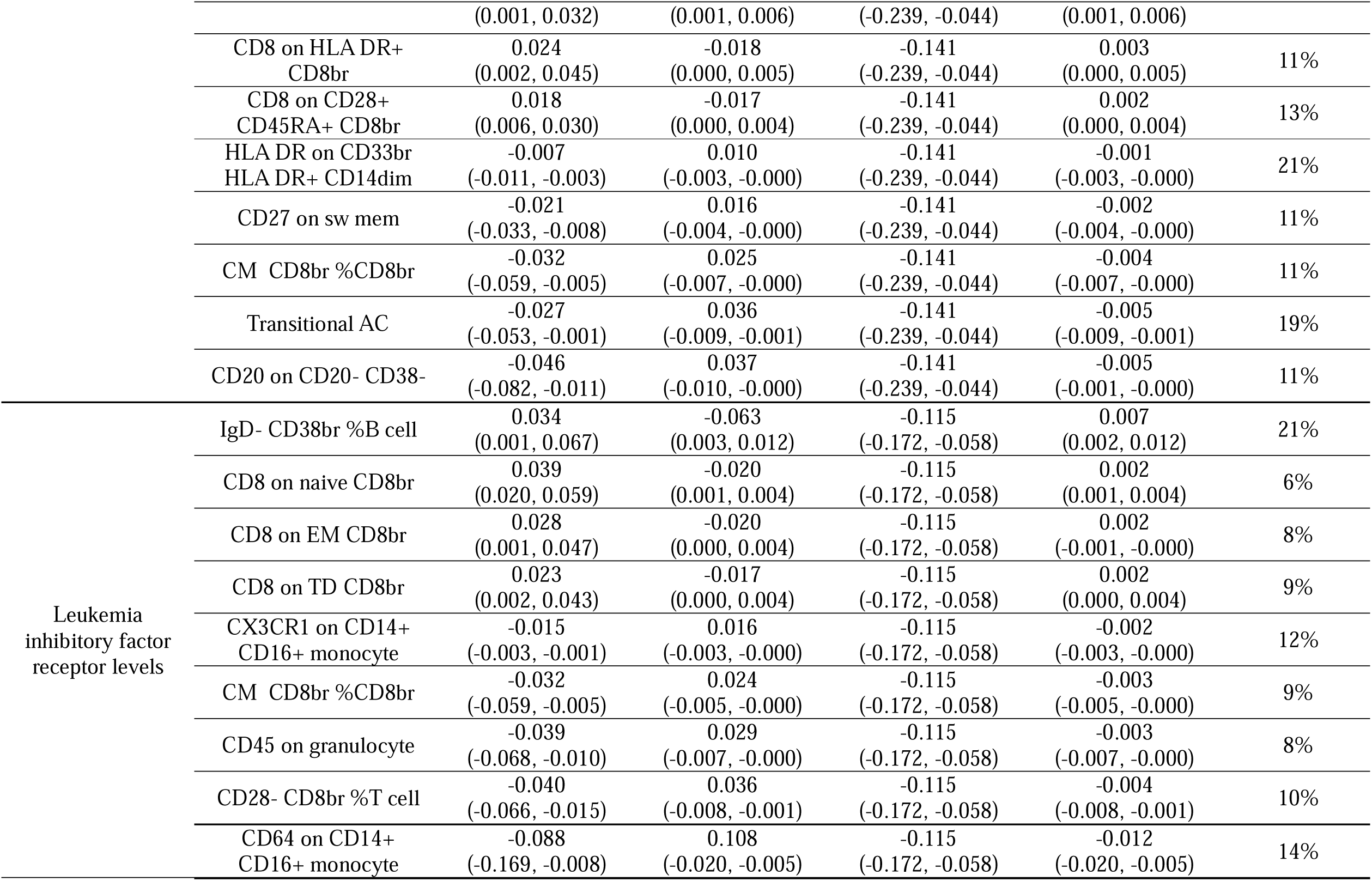
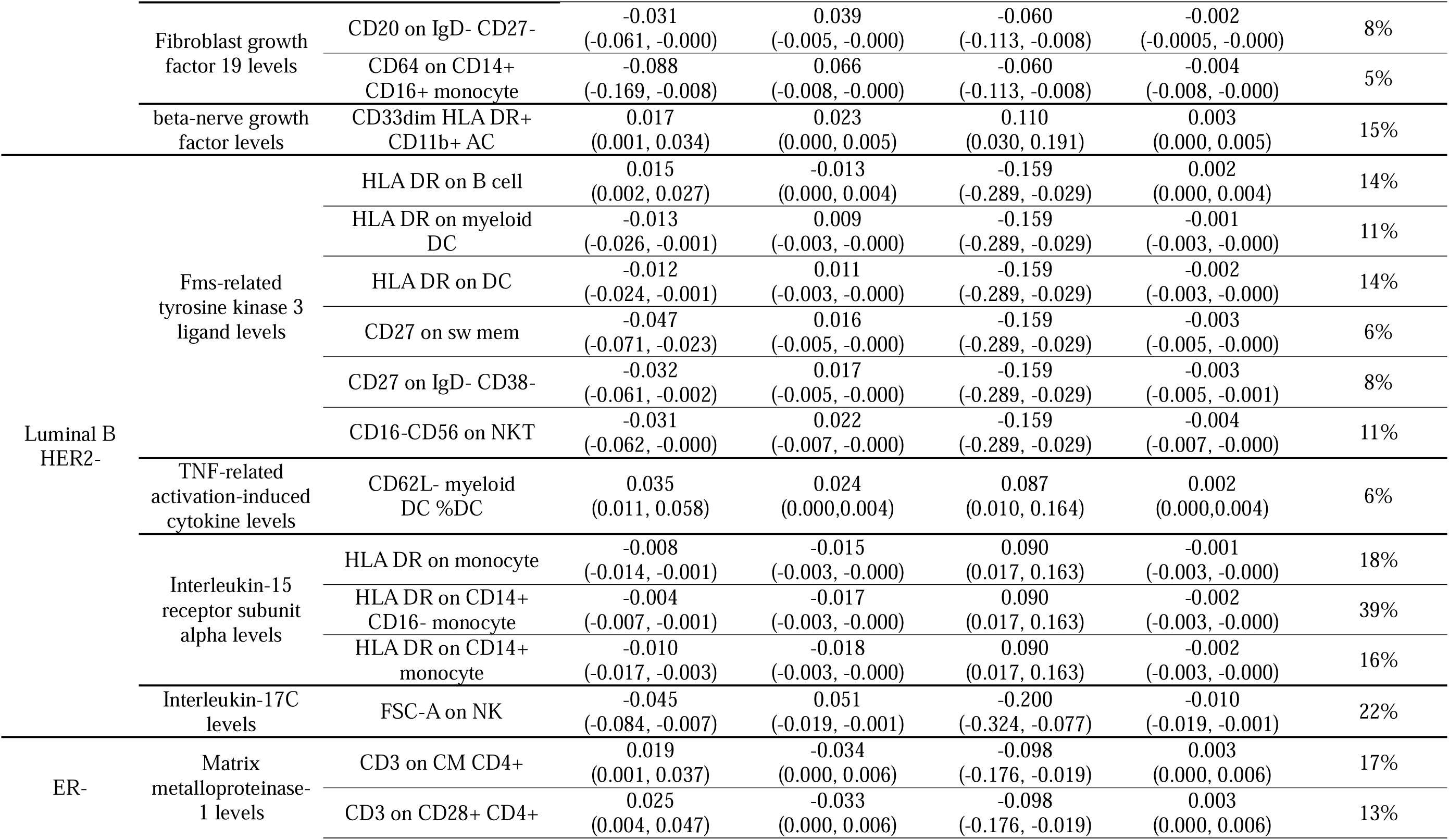
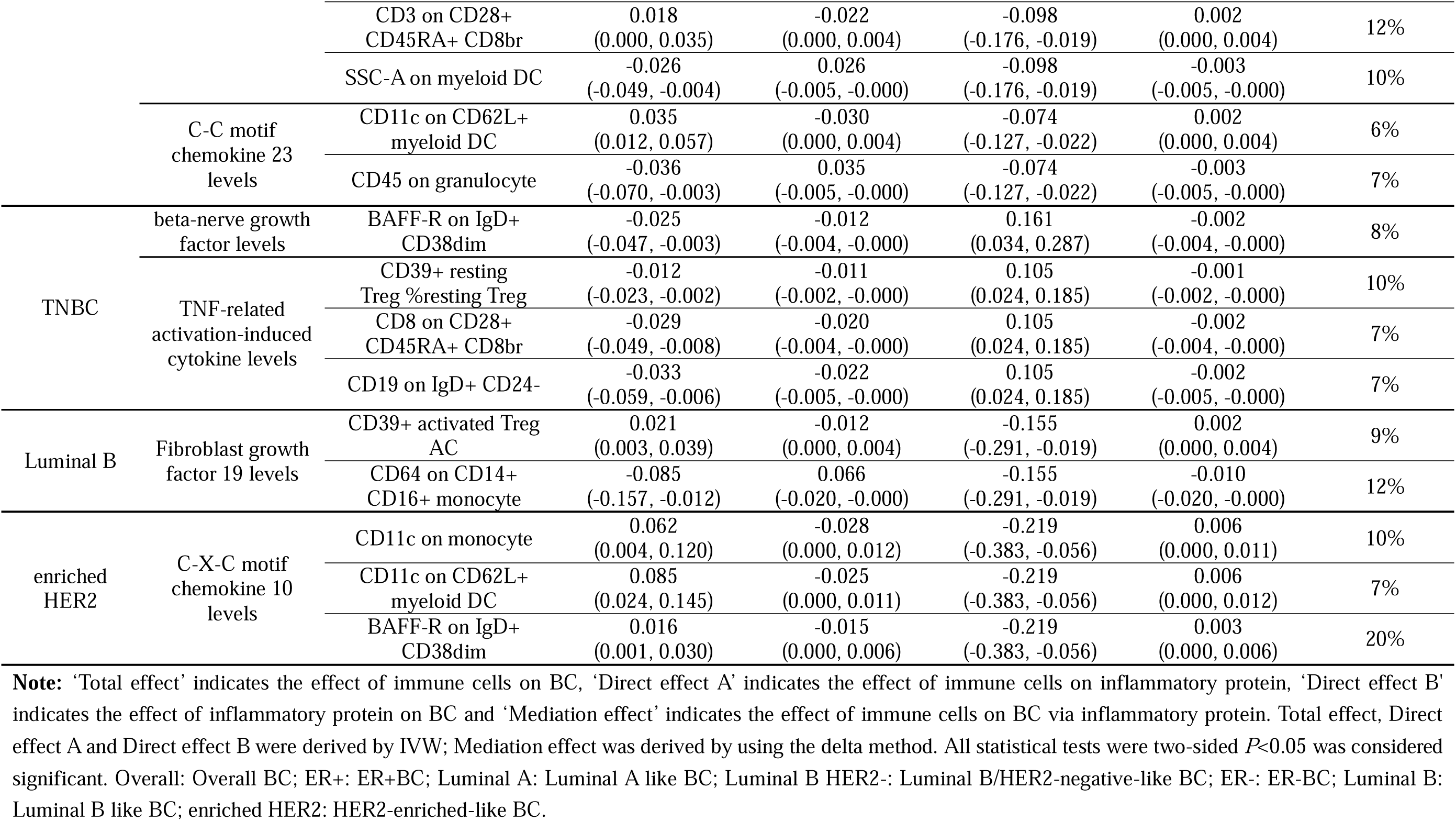
The mediation effect of immune cells on BC via inflammatory protein.

## 4. Discussion

To the best of our knowledge, this is the first study to comprehensively evaluate the causal relationships among immune characteristics, inflammatory protein, and BC. In this study, a bi-directional two-sample MR analysis was employed to thoroughly investigate the causal associations between immune cells and BC and its subtypes. Additionally, we systematically examined the mediating effects of inflammatory proteins on the relationship between immune cells and BC. After using FDR correction, sixteen immune traits were found to have a significantly causal association with the risk of BC, indicating their potential role in the development of BC. This study not only enhances our comprehension of the immunological landscape of BC but also provides vital evidence for developing more precise and personalized treatments.

Previous studies have demonstrated the association between immune traits and BC by using MR methods [37, 38]. Findings indicate discrepancies with previous conclusions regarding ER-, ER+, and overall breast cancer, suggesting these differences may arise from variations in the explanatory power of selected genetic variants (kb) and the correlation between instrumental variables and phenotypes (r^2^) [39]. This highlights the critical importance of meticulously considering the explanatory power of genetic variants and their correlation with phenotypes to ensure consistency and reliability in MR analysis outcomes. However, this study uses the MR approach more systematically to explore the specific associations between BC molecular subtypes (such as Luminal A-like, Luminal B-like, and Luminal B/HER2-negative-like BC) and immune traits, an aspect not explored in previous MR research and reveals the heterogeneity of immune cells within BC molecular subtypes. In addition, research by Chung et al. using single-cell RNA sequencing also highlights the importance of immune cell heterogeneity in BC for the complex interactions between the tumor and the immune system [40]. The diverse immune cell landscape is crucial for the development of targeted treatment strategies, as the interactions between tumor cell populations and their microenvironment directly determine treatment outcomes and drug resistance mechanisms [41]. Therefore, it is essential to systematically investigate the role of different immune traits in different BC subtypes by using MR analysis. By in-depth analysis of specific immune traits in different BC molecular subtypes, this study illustrates the potential mechanisms of specific BC molecular subtypes and highlights the complexity and importance of the immune microenvironment in cancer development.

In this study, HLA DR on monocytes was found to be associated with a decreased risk of luminal A-like BC and BRCA1-mutated TNBC (**Figure 2** and **Table 3)**, with various cell subpopulations expressing HLA DR conferring a protective effect against BRCA1-mutated TNBC (**Figure 2** and **Table 3)**. Specifically, HLA DR on B cell, located on TBNK, was associated with an increased risk of HER2-enriched-like BC (**Figure 2** and **Table 3**), in contrast to its role in other BC subtypes. Our results demonstrate that high expression of HLA-DR may play different important roles in different breast cancer subtypes. Human leukocyte antigen-DR (HLA-DR), a type of MHC II molecule, is primarily expressed on professional antigen-presenting cells (APCs) and is considered critical for antigen presentation [42]. The expression of HLA-DR on monocytes is significant for tumor development, as tumors may evade immune responses by suppressing HLA-DR expression. Conversely, therapeutic strategies aimed at increasing HLA-DR expression may enhance anti-tumor immunity. Existing research confirms that high expression of HLA-DR is closely associated with high expression of CD4 and ICOS in the breast tumor microenvironment, underscoring the indispensable role of HLA-DR in the activation of CD4+ T cells, thereby promoting the hypothesis of enhancing host anti-tumor immune responses [43]. In addition, studies have shown that high expression of HLA class II genes may to some extent enhance the visibility of tumor cells to the immune system, promote effective immune responses, and be associated with improved prognosis[44]. These pieces of evidence support the key function of HLA-DR in the immune battle against BC, which is consistent with our observations. Similar conclusions have been drawn in TNBC, where the expression of HLA-DR in TNBC plays a critical role in favorable patient prognosis [45]. However, the expression regulation of class II MHC molecule genes shows different patterns in various tumor immune microenvironments. Studies indicate that IFN-_γ_ can induce high levels of class II MHC molecules in tumor cells, which is associated with malignant progression and beneficial for tumor cell survival and proliferation [46]. Therefore, we propose that high levels of IFN-_γ_ in HER2-enriched-like BC may promote high expression of HLA-DR on B cells, facilitating tumor cell survival and proliferation, leading to its different role in BC. Thus, the specific role of HLA-DR in BC development is complex and multifaceted, and further research is needed to elucidate its role in the development of different BC molecular subtypes.

In this study, four types of B cells with CD25 expression, including CD25 on CD24+ CD27+, CD25 on IgD- CD38dim, CD25 on memory B cell, and CD25 on sw mem, were confirmed to have a significant protective effect against HER2-enriched-like BC (**Figure 2** and **Table 3**). Our research indicates that high expression of CD25 on B cells is an important protective factor against BC. However, CD25 is typically highly expressed on activated T cells (including Tregs), which can infiltrate tumor tissues and suppress anti-tumor immune responses, thereby promoting cancer progression [47].In B cells, CD25 expression is located in the CD27+ memory cell subpopulation. These CD25+ B cells have efficient antigen-presenting capabilities and can secrete higher levels of the immunoregulatory cytokine IL-10, suggesting that they may function as a unique immunoregulatory B cell population that plays a role in autoimmune diseases[48]. Another study showed that the transcriptional characteristics of memory B cells are associated with improved overall and disease-free survival in TNBC patients [49]. In addition, IL-10 secretion has been shown in mouse models to promote the proliferation and effector functions of CAR-T cells, enabling complete elimination of solid tumors and metastatic cancers, including multiple cancers such as colon, breast, melanoma and pancreatic cancer [50]. However, there is currently no evidence to suggest a direct relationship between CD25+ B cells and BC. It is possible that CD25-expressing B cells may play a protective role in BC, but further investigation is needed to determine their precise functions and roles in the disease.

This study confirms that CD27 on sw mem has a protective role against both Luminal B/HER2-negative-like BC and Luminal A-like BC (**Figure 2** and **Table 3**). Elevated levels of CD27 on IgD- CD38dim B cells also reduce the risk of Luminal B/HER2-negative-like BC (**Figure 2** and **Table 3**). Therefore, it is posited that CD27 serves a protective function across multiple BC subtypes.CD27, also referred to as TNFRSF7, is a type I transmembrane glycoprotein that, when bound to its ligand CD27L (CD70), participates in signaling pathways critical for the regulation of cellular functions [51, 52]. The efficacy of antibodies that target the CD27/CD70 axis has been validated in preclinical models of lymphoma, renal cell carcinoma (RCC), breast cancer, and sarcoma, highlighting their potential in cancer therapy [53]. Research has emphasized the vital role of CD27 in enhancing immune cell infiltration, T cell differentiation, and macrophage polarization, all of which are essential for anti-tumor immune responses [54]. Specifically, a unique subset of B cells expressing pro-inflammatory markers has been identified in BC patients. These cells are characterized by CD27 and other markers (e.g., IgG+ excluding IgM or IgD) and have been shown to promote anti-tumor T cell responses [55]. This suggests that CD27+ B cells in the BC microenvironment may play a critical role in enhancing the immune system’s ability to fight tumors, supporting our findings. Targeting these B cell subsets may offer new immunotherapeutic avenues for BC treatment.

In reverse MR analysis, a significant association was confirmed between TNBC and reduced CD3 on TD CD4+, indicating that the presence of TNBC leads to a decrease in CD3 on TD CD4+ (**Figure 5** and **Supplementary Table S5**). Previous studies have shown that PD-L1 is expressed in approximately 20% of TNBC cases [56]. The activation of PD-1, mediated by its ligands PD-L1 or PD-L2, inhibits the signaling cascade triggered by the CD3/TCR complex. This leads to a reduction in the production of IL-2, attenuation of T-cell proliferation, and a decrease in cytotoxic activity [57]. In cancer therapy, PD-1 or PD-L1 inhibitors can be used to block the PD-1 pathway, removing the inhibition of CD3/TCR signaling and restoring T-cell activation, thereby regaining the ability of T-cells to recognize and kill tumor cells.

In this study, CD40L receptor levels were identified as significant mediators of the causal relationship between immune cells and the BC (**Table 4**). Previous observational studies have confirmed that CD40L receptor levels are significantly higher in both B and T cells in BC patients [58]. Furthermore, CD40/CD40L connection plays a dual role in tumor biology: membrane-bound CD40L strongly activates signaling and may eradicate tumors, while soluble CD40L, released primarily from activated T lymphocytes and platelets, can inhibit apoptosis [59]. lymphocytes and surface-released soluble CD40L from activated platelets help tumor survival by inhibiting apoptosis, suppressing the immune system, and promoting angiogenesis [60]. These findings are consistent with our results, which indicate that CD40L can either increase or decrease the risk of BC.

This study has limitations that cannot be overlooked. First, MR analysis cannot substitute for clinical trials in objective domains, as it only analyzes causal associations between exposure and outcome. Therefore, further research is needed to confirm the potential association between immune traits and BC risk. On the other hand, due to the limited availability of GWAS data resources, our MR analysis was conducted solely in European populations. Because of the genetic heterogeneity among different ethnic groups, our study results cannot be generalized to other populations. Furthermore, the lack of individual information prevented us from conducting further stratified analyses. Consequently, future research should include stratified analyses of different populations to draw more comprehensive conclusions. Lastly, our study utilized a more relaxed threshold to evaluate the results, which may increase the occurrence of some false positives, while allowing a more comprehensive evaluation of the strong associations between immune traits and BC.

## 6. Conclusions

This study significantly predicts causal associations between 16 immune traits and the overall risk of BC and its subtypes, as well as 108 causal associations mediated by inflammatory proteins. The 16 causal associations include CD25 on CD24+ CD27+, CD25 on IgD- CD38dim, CD25 on memory B cells, CD25 on switched memory (sw mem), CD27 on IgD- CD38dim, CD27 on switched memory (sw mem), CD40 on CD14-CD16+ monocytes, CD8 on naive CD8br, DC AC, HLA DR on B cells, HLA DR on CD14+ CD16-monocytes, HLA DR on CD14+ monocytes, HLA DR on CD33br HLA DR+ CD14dim, HLA DR on monocytes, and HLA DR++ monocyte %monocyte. Mediation analysis revealed that CD40L receptor levels mediate up to 73% of the relationship between IgD- CD27-%B cells and ER+BC. These markers could unveil novel drug targets for combating breast cancer. Though additional experimental research is necessary to thoroughly investigate the potential roles of immune traits in the pathophysiology of both breast cancer overall and its molecular subtypes, this study provides potential predictive markers for personalized immunotherapy in BC patients, offering important evidence to guide clinical treatment decisions.

## Abbreviations

BC: Breast cancer
BCAC: Breast Cancer Association Consortium
CIMBA: Consortium of Investigators of Modifiers of BRCA1/2
DRIVE: Discovery, Biology and Risk of Inherited Variants in Breast Cancer Consortium
ER-: Estrogen Negative
ER+: Estrogen Positive
TILs: Tumor Infiltrating Lymphocytes
TNBC: Triple-negative breast cancer
MDSC: Myeloid-derived suppressor cells
MR: Mendelian Randomization
IVs: Instrumental variables
IVW: Inverse Variance Weighted
GWAS: Genome-Wide Association Studies
AC: Absolute Cell
MFI: Median Fluorescence Intensities
MP: Morphological Parameters
FSC: forward scatter
SSC: side scatter
RC: Relative Cell
SNPs: single nucleotide polymorphisms
ER: estrogen receptor
PR: progesterone receptor
HER2: human epidermal growth factor receptor 2
LD: linkage disequilibrium
OR: odds ratio
CI: confidence intervals
FDR: False Discovery Rate
HLA: Human leukocyte antigen

## Supplementary Information Supplementary Table

**Table S1.** The instrumental variables for immune traits used in MR analyses. **Table S2.** Full result of MR estimates for the association between Immune cells and Breast Cancer. **Table S3.** Sensitivity analysis results of the significantly association between Breast Cancer and immune cells. **Table S4.** The instrumental variables for Breast cancer used in MR analyses. **Table S5**. Full result of MR estimates for the association between Breast Cancer and Immune cells. **Table S6.** Sensitivity analysis results of the significantly association between immune cells and Breast Cancer. **Table S7**. The mediation effect of immune cells on Breast Cancer and Subtypes via Inflammatory Protein. **Table S8**. The instrumental variables for Inflammatory Protein used in MR analyses. **Table S9**. Full result of MR estimates for the association between inflammatory protein on Breast Cancer and Subtypes.

## Declarations

### Ethics Approval and Consent to Participate

This study employs publicly available summary-level data; therefore, no ethics approval was required, and informed consent was not applicable. Our research conforms to all relevant guidelines for the use of such data.

### Consent for Publication

Not applicable.

### Availability of Data and Materials

Publicly available datasets were analyzed in this study. The GWAS summary statistics for 731 immune traits could be publicly available in the GWAS Catalog (https://www.ebi.ac.uk/gwas/). Summary statistics for breast cancer and its subtypes are available at http://bcac.ccge.medschl.cam.ac.uk/bcacdata/.

### Declaration of Interest Statement

The authors declare that they have no competing interests.

## Funding

This research was funded by Talent Development Foundation of The First Dongguan Affiliated Hospital of Guangdong Medical University (PF100-2-02), and the Key Discipline Construction Project of Guangdong Medical University (Grant No. 4SG23004G).

## Authors’ Contributions

Conceptualization and Design: QG, JY, YW, and ZH; Data curation and Formal Analysis: QG, JY, and YD; Visualization: QG and JY; Writing – original draft: QG, JY, and YW; Writing – review & editing: ZH, AF, and KZ; Supervision and Funding acquisition: ZH. All authors contributed to the article and approved the submitted version.

## Acknowledgments

Not applicable.

## Supplementary Figure

**Figure S1.**
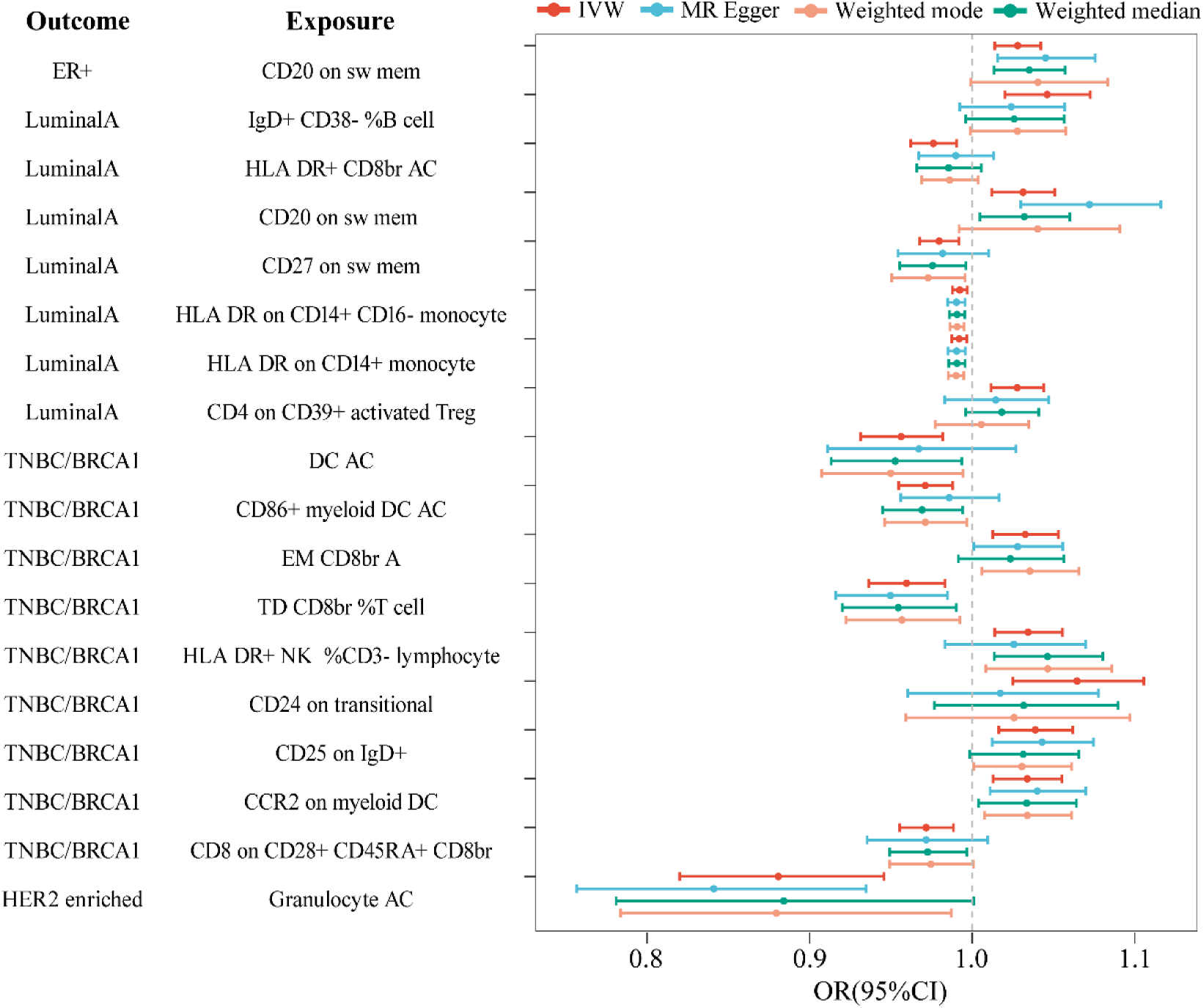
Forest plot revealing the suggestive association between immune traits and breast cancer and its molecular subtypes through different approaches. **Note:** CI: Confidence Interval; OR: Odds Ratio; ER+: ER+BC; Luminal A: Luminal A like BC; TNBC/BCRA1: BRCA1-mutated TNBC; HER2 enriched: HER2-enriched-like BC.

**Figure S2.**
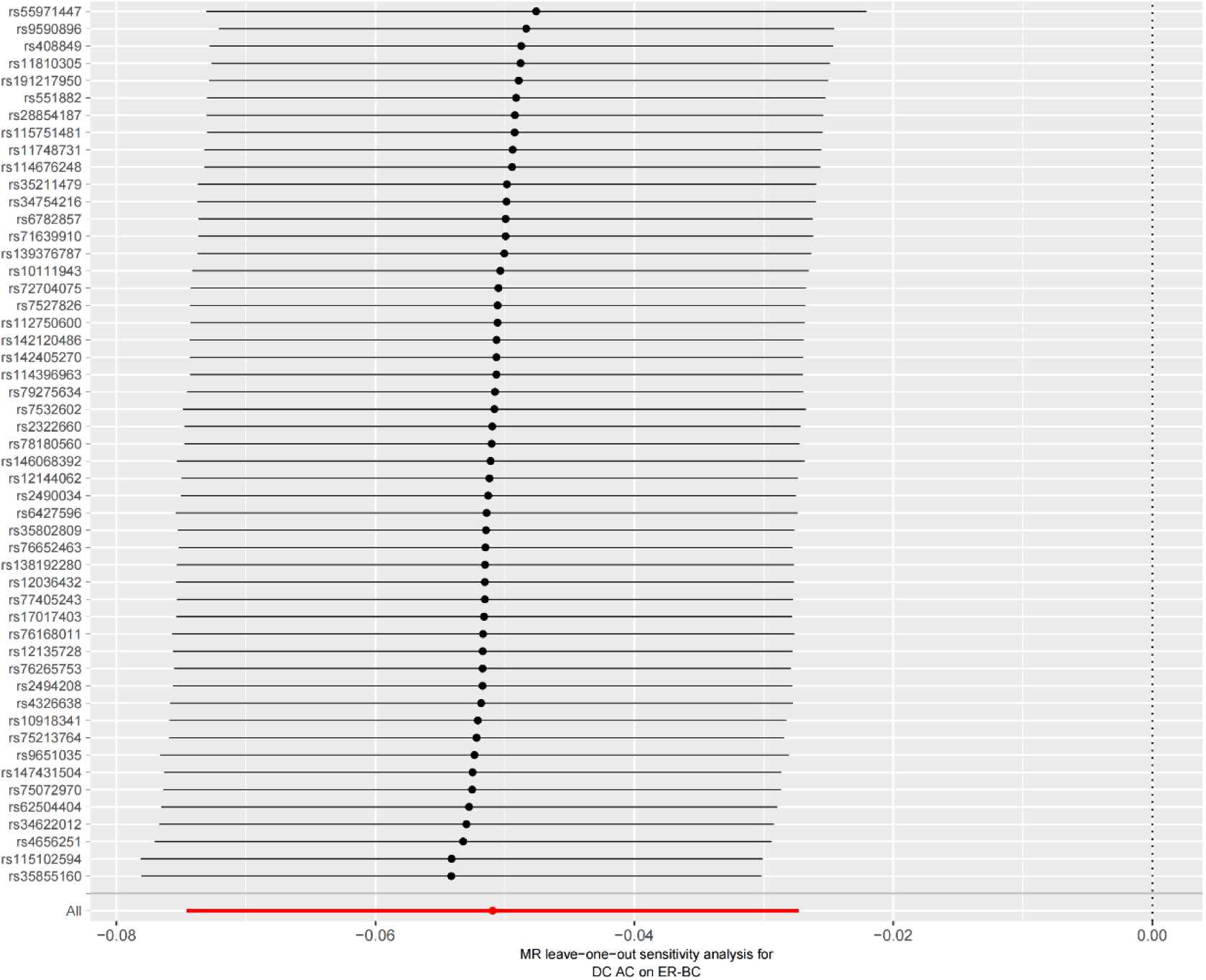
Leave-one-out forest plot of a two sample Mendelian randomization study of immune traits (DC AC) and ER-defined typing of BC (ER-BC).

**Figure S3.**
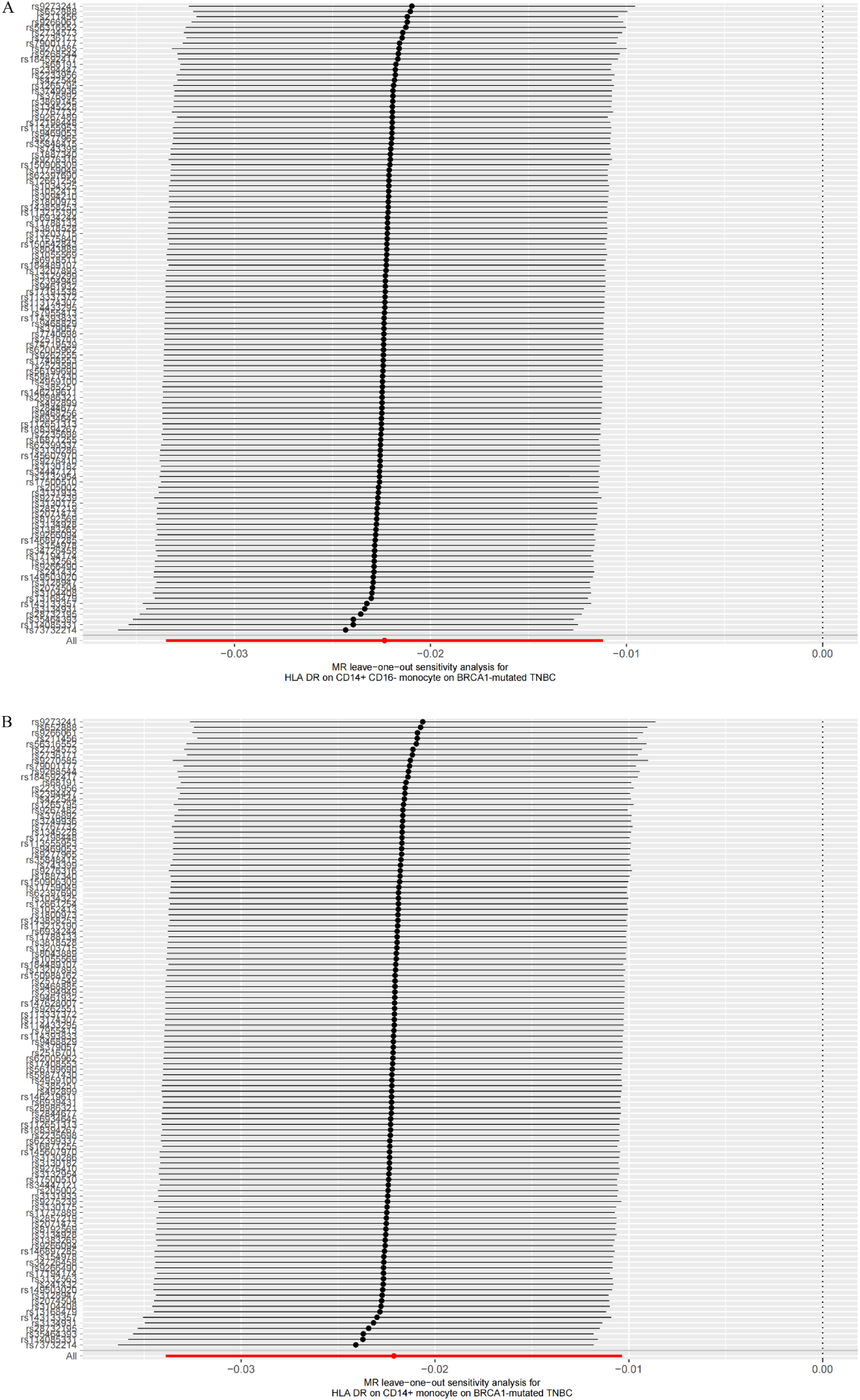
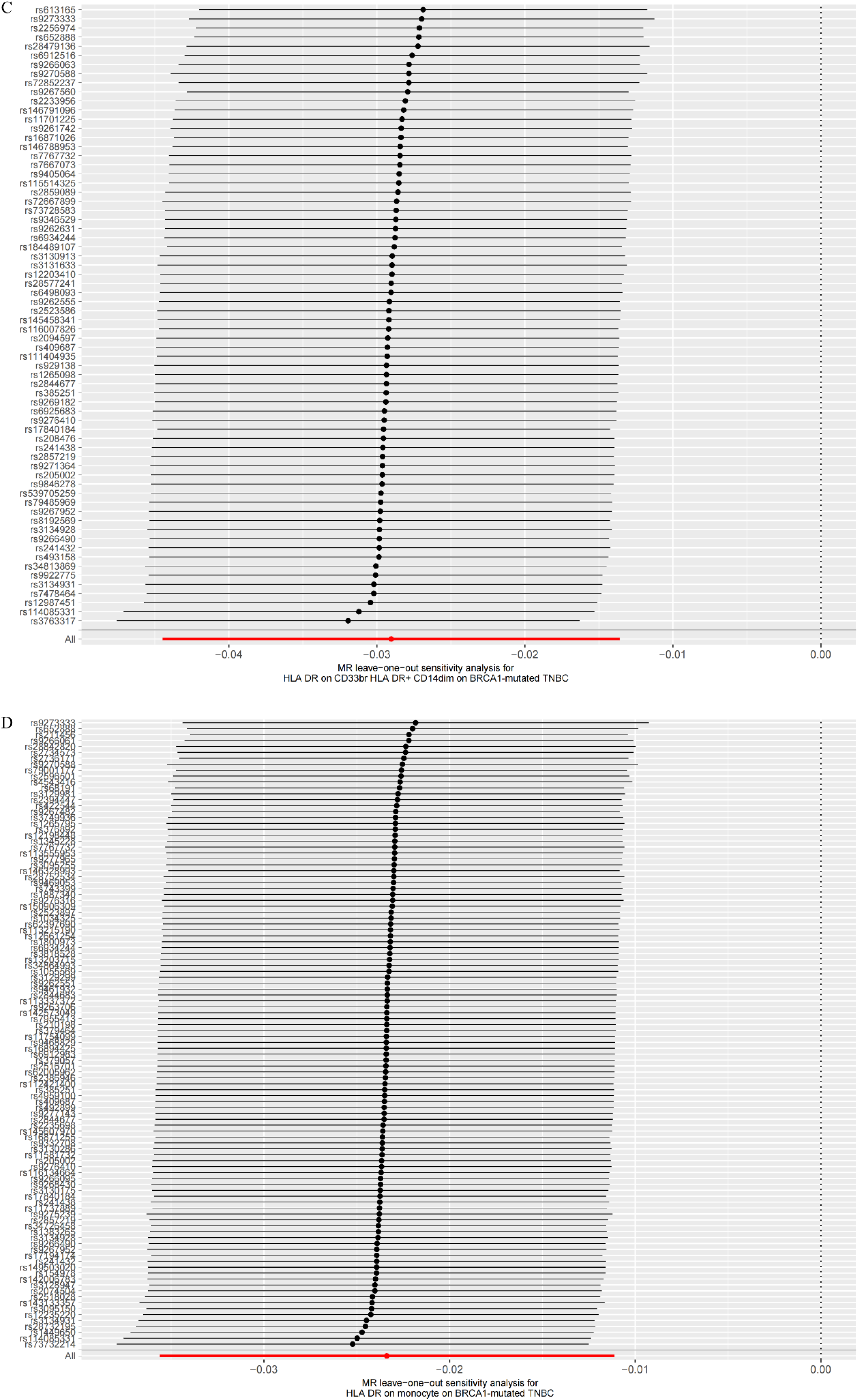
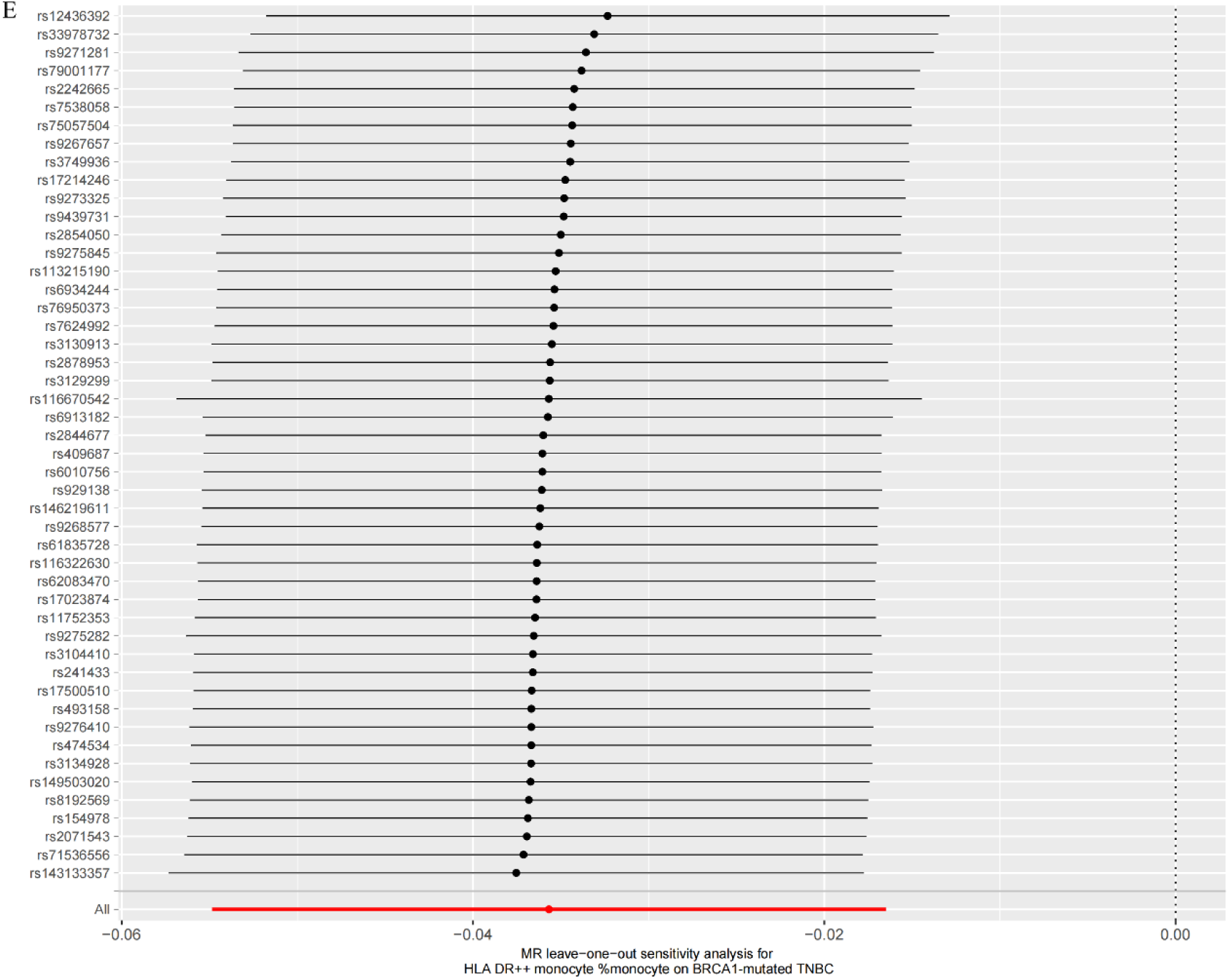
Leave-one-out forest plot of a two sample Mendelian randomization study of immune traits and BRCA1-mutated TNBC. **(A)** HLA DR on CD14+ CD16-monocyte on BRCA1-mutated TNBC. **(B)** HLA DR on CD14+ monocyte on BRCA1-mutated TNBC. **(C)** HLA DR on CD33br HLA DR+ CD14dim on BRCA1-mutated TNBC. **(D)** HLA DR on monocyte on BRCA1-mutated TNBC. **(E)** HLA DR++ monocyte %monocyte on BRCA1-mutated TNBC.

**Figure S4.**
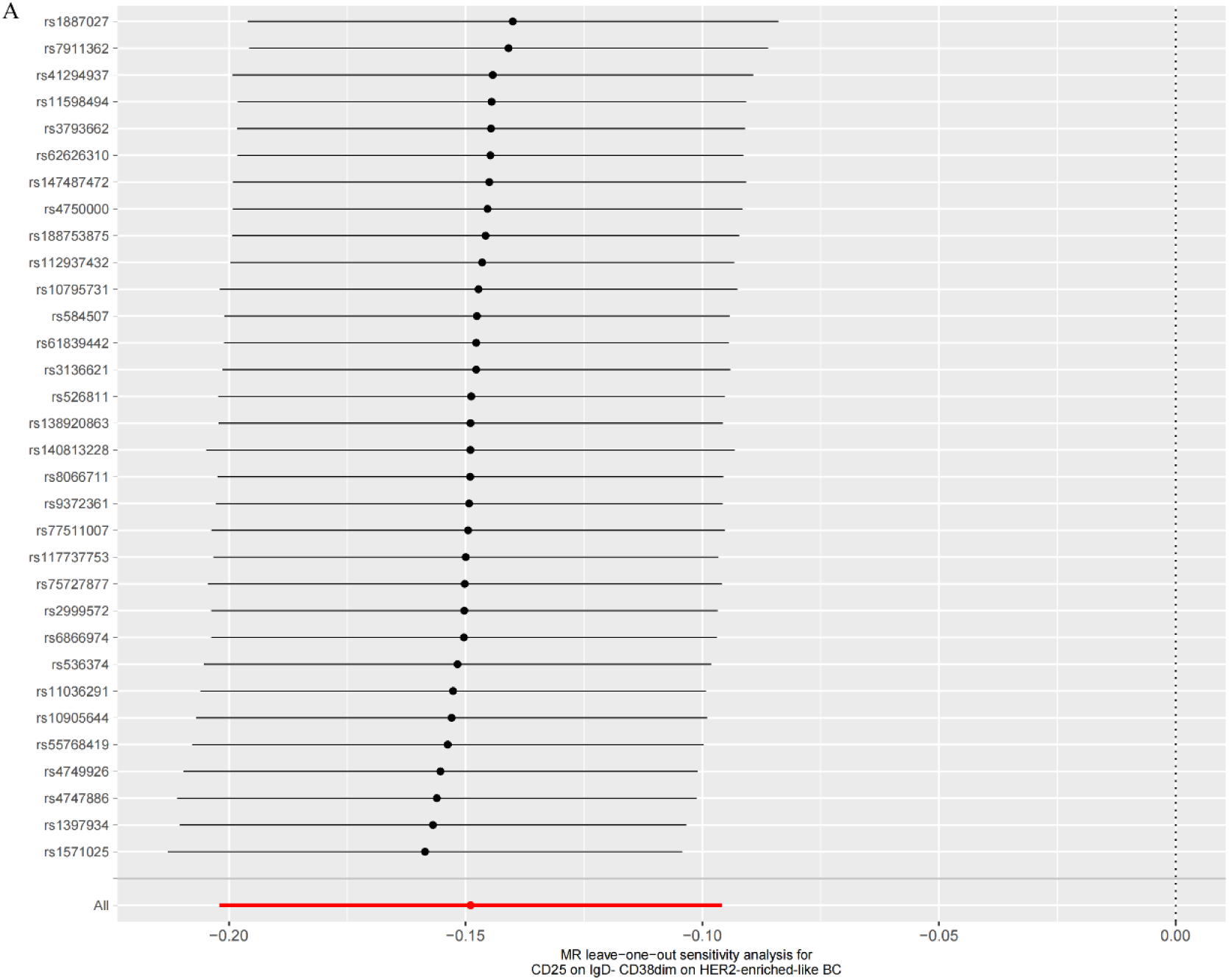
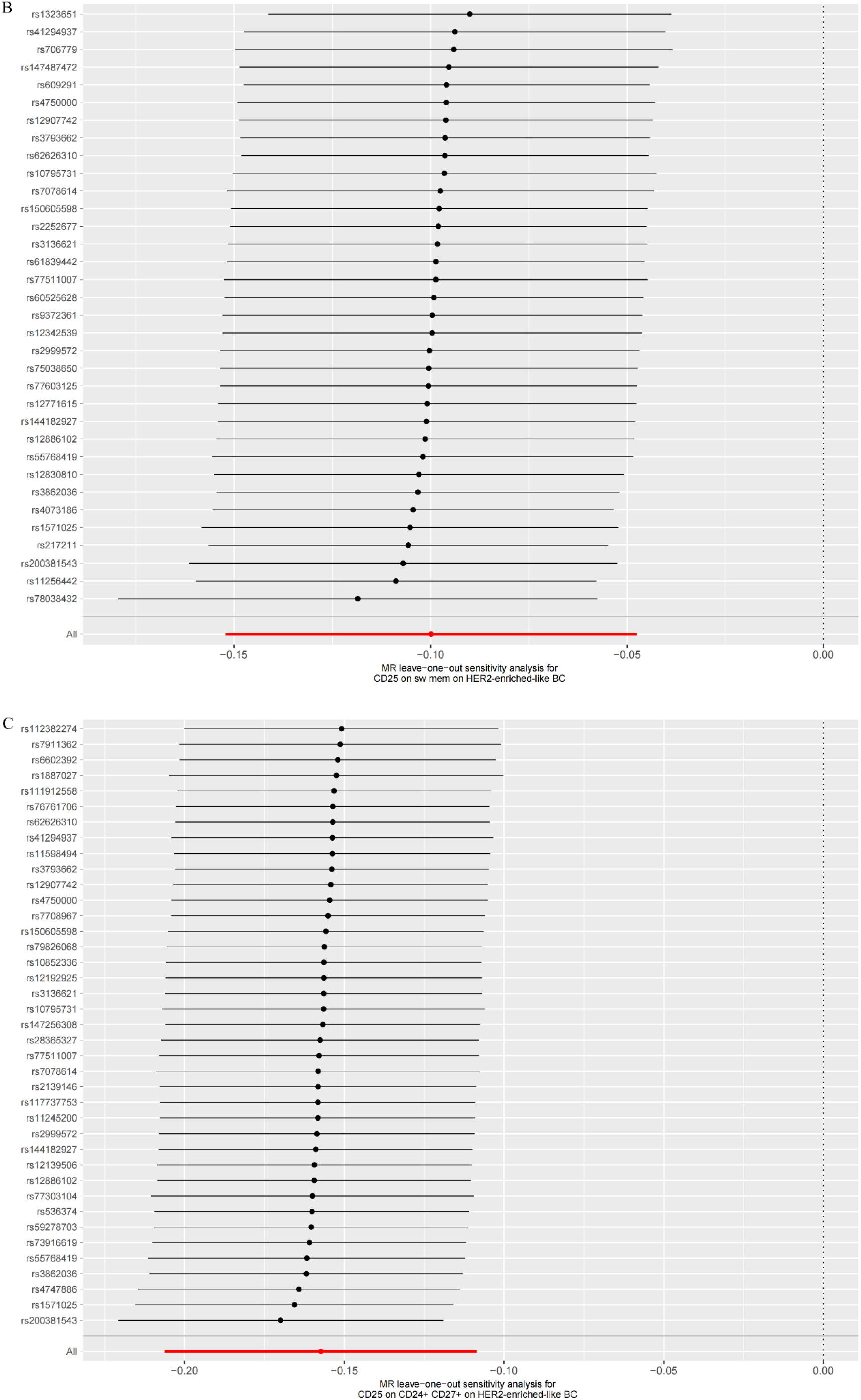
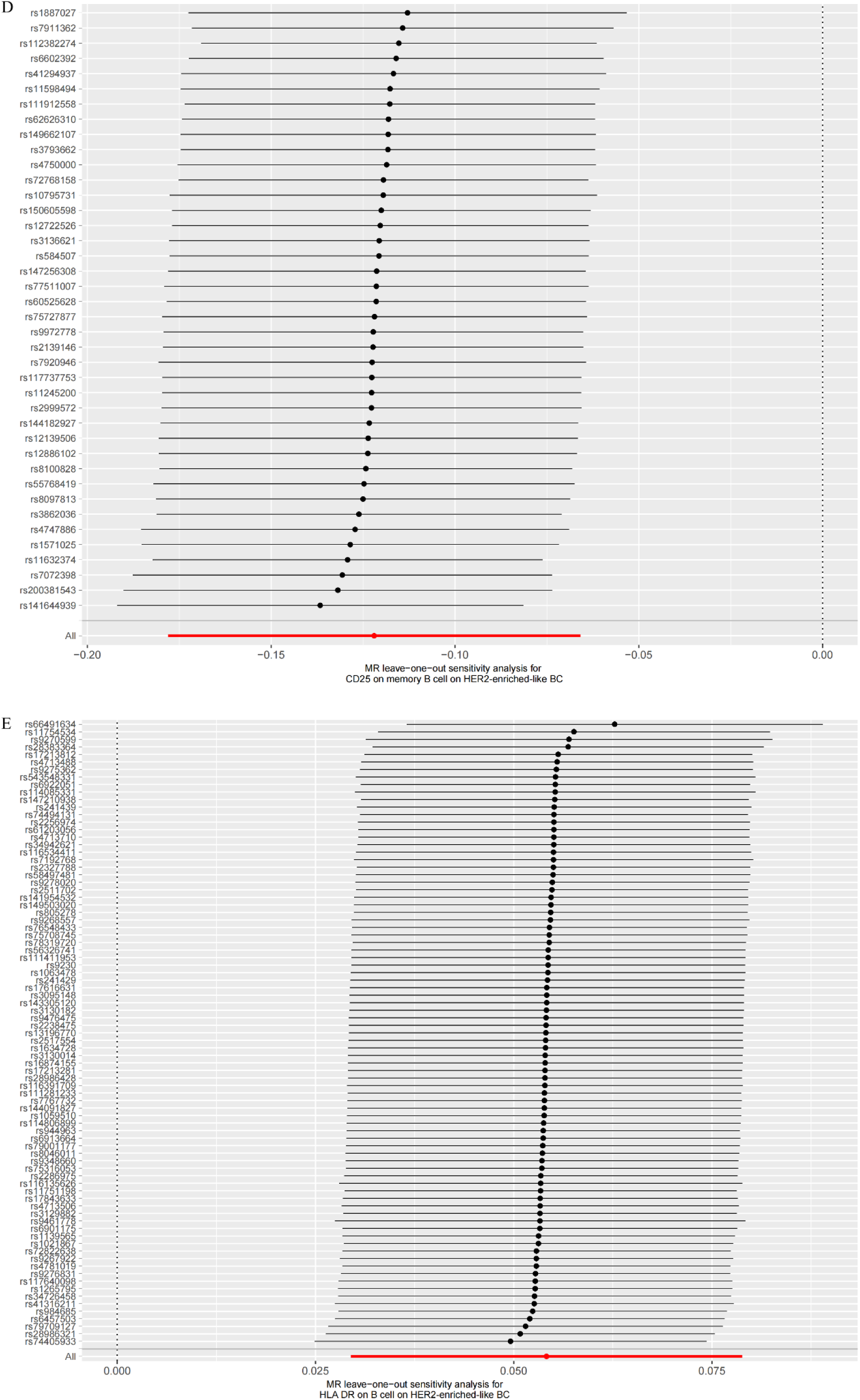
Leave-one-out forest plot of a two sample Mendelian randomization study of immune traits and HER2-enriched-like BC. (A) CD25 on IgD- CD38dim on HER2-enriched-like BC. (B) CD25 on sw mem on HER2-enriched-like BC. (C) CD25 on CD24+ CD27+ on HER2-enriched-like BC. (D) CD25 on memory B cell on HER2- enriched-like BC. (E) HLA DR on B cell on HER2-enriched-like BC

**Figure S5.**
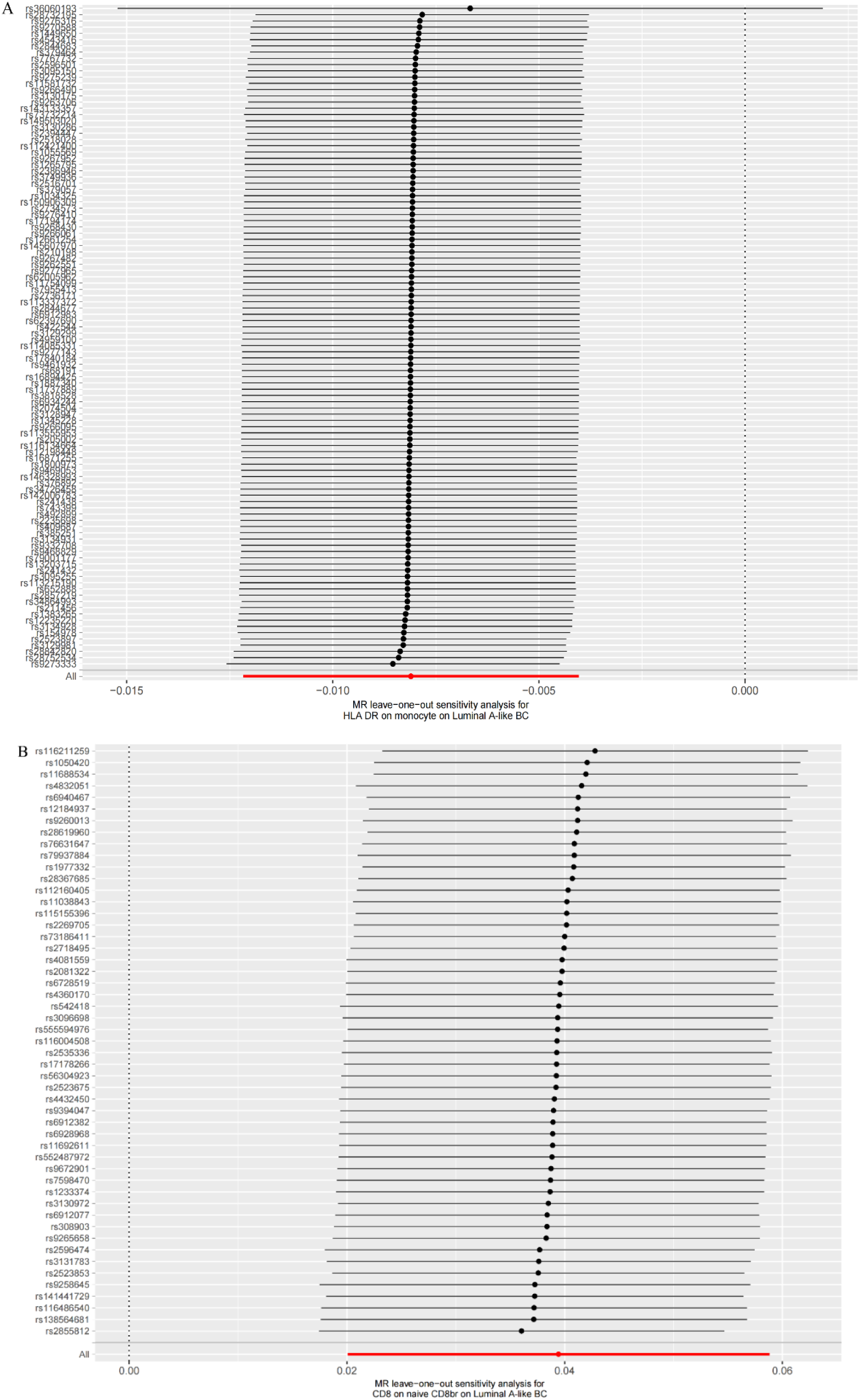
Leave-one-out forest plot of a two sample Mendelian randomization study of immune traits and Luminal A. (A) HLA DR on monocyte on Luminal A-like BC. (B) CD8 on naive CD8br on Luminal A-like BC.

**Figure S6.**
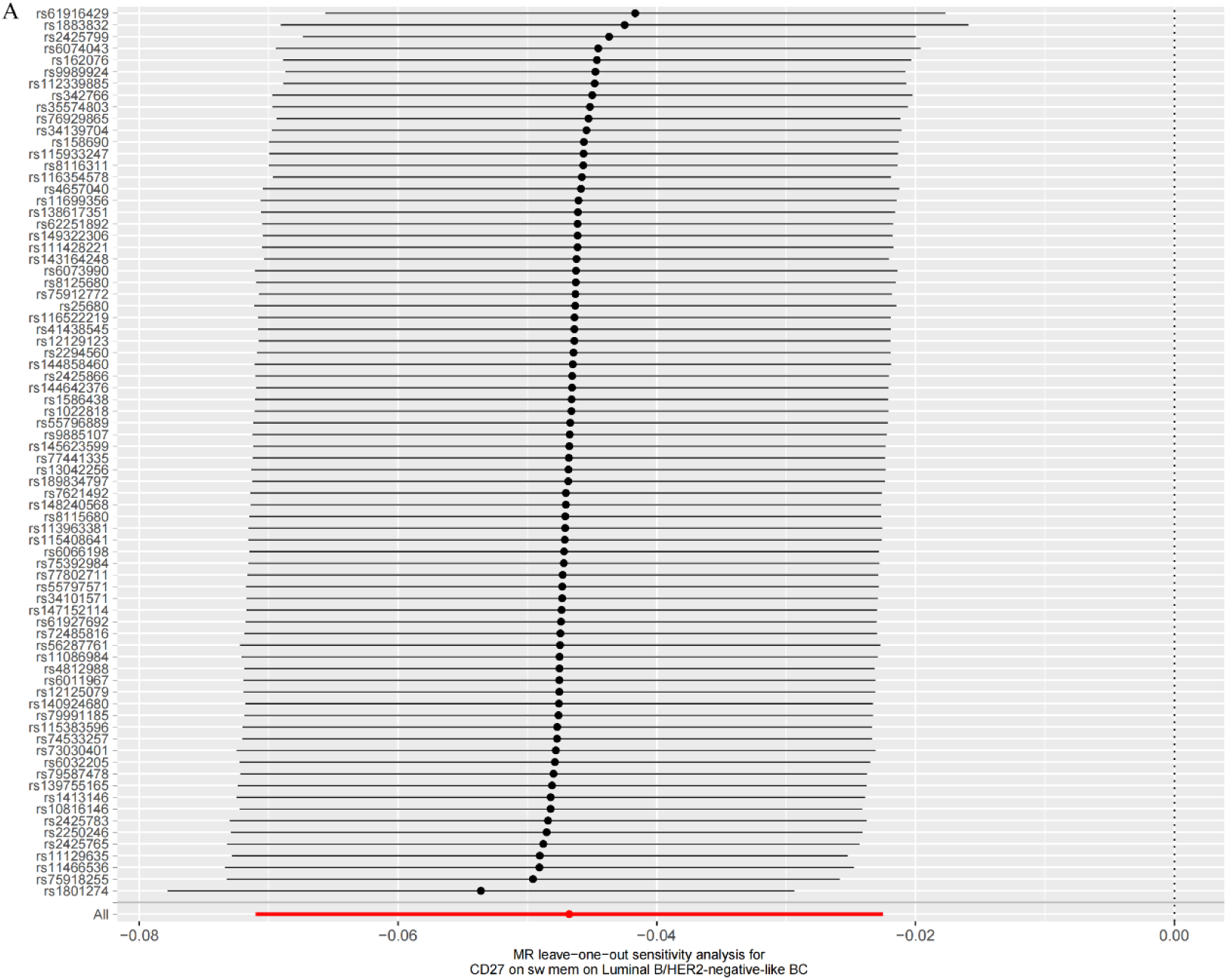
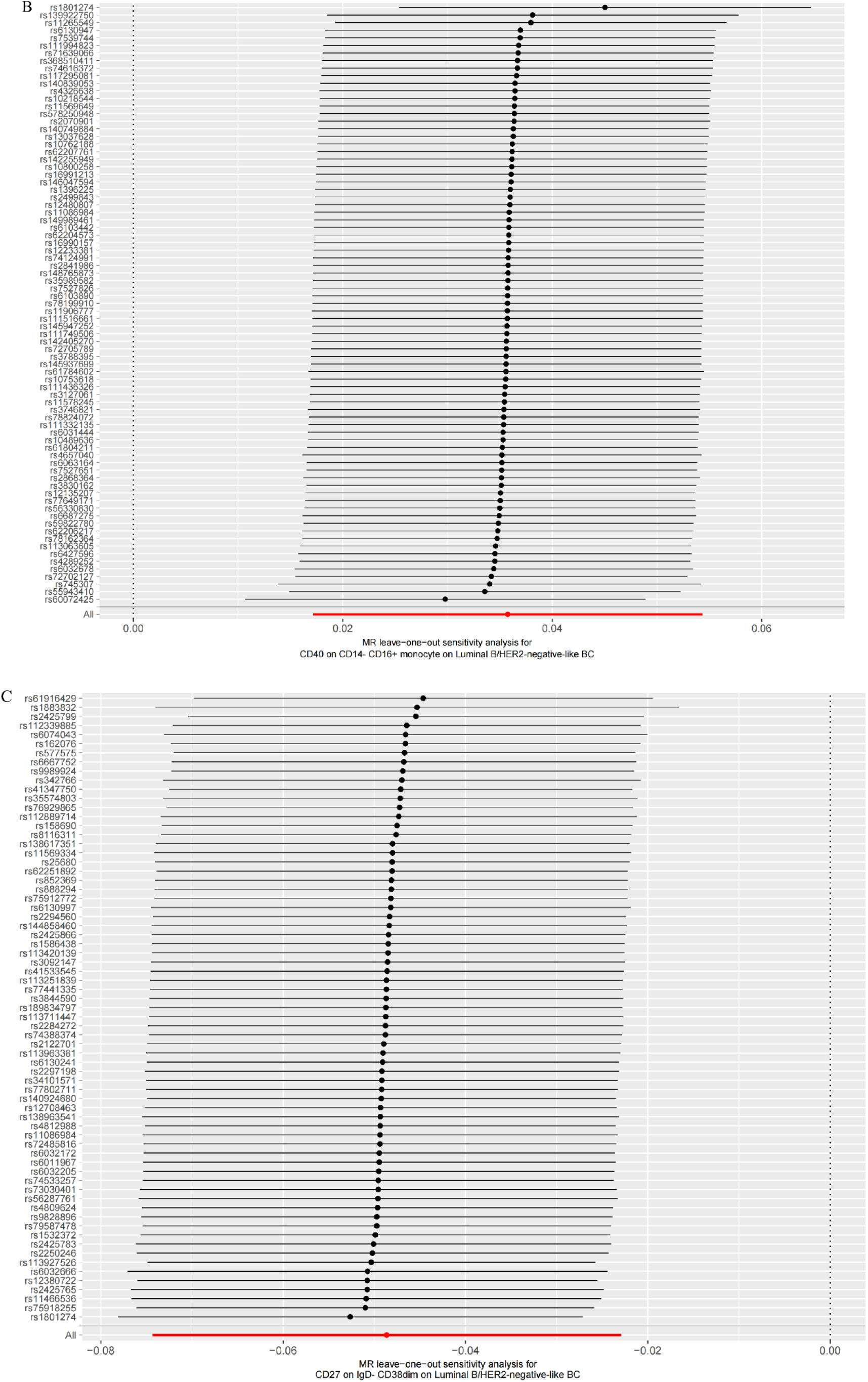
Leave-one-out forest plot of a two sample Mendelian randomization study of immune traits and Luminal B HER2-. (A) CD27 on sw mem on Luminal B/HER2- negative-like BC. (B) CD40 on CD14- CD16+ monocyte on Luminal B/HER2- negative-like BC. (C) CD27 on IgD- CD38dim on Luminal B/HER2-negative-like BC

**Figure S7.**
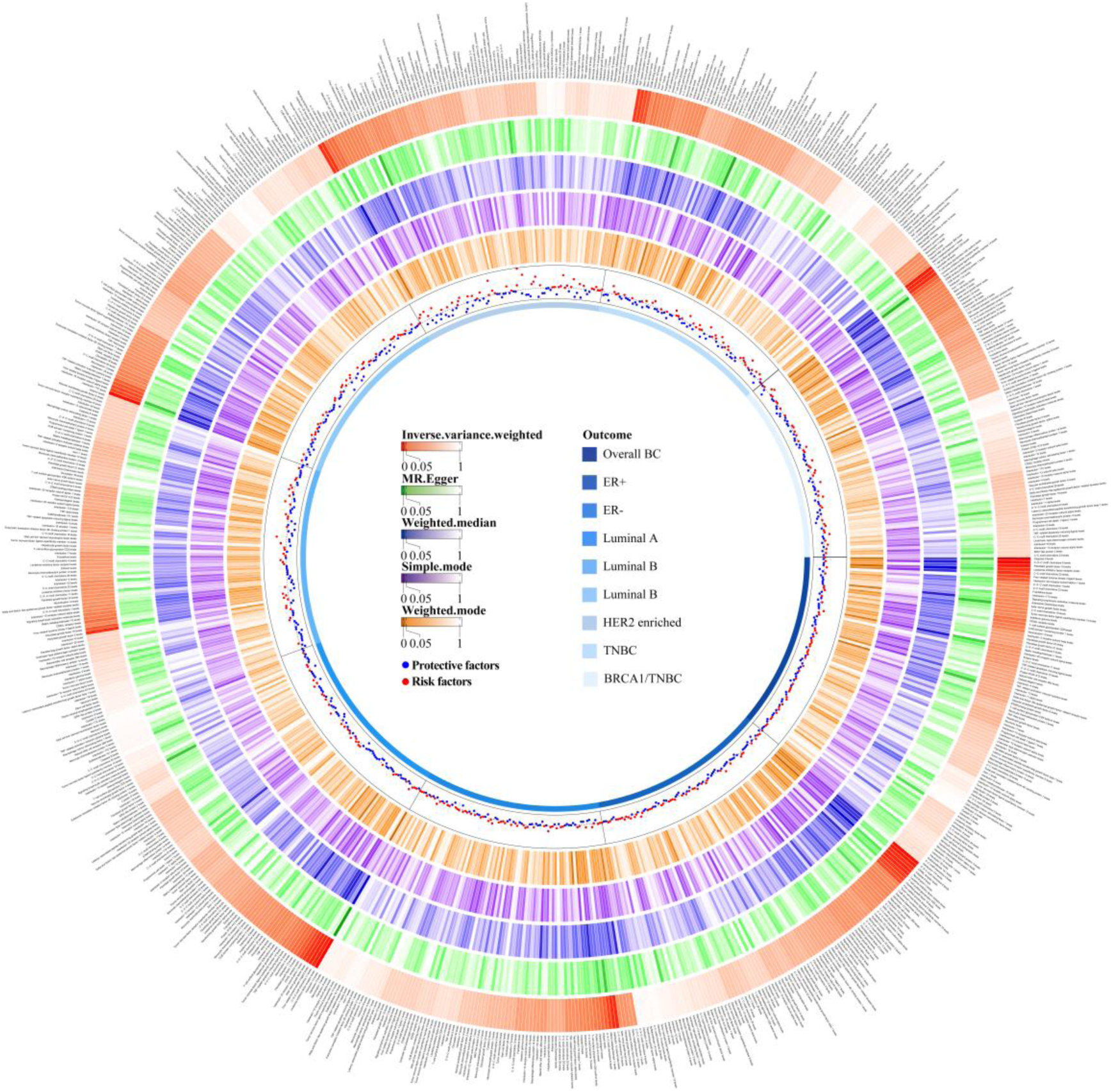
Heatmap depicting p-values for inflammatory proteins and breast cancer for all results: the outer circle indicates the name of the inflammatory proteins, the middle circle indicates the p-values for the results of the different MR analysis methods in different colors, the inner circle indicates the breast cancer and its subtypes, and the red and blue dots indicate the direction of the odds ratios

**Figure S8.**
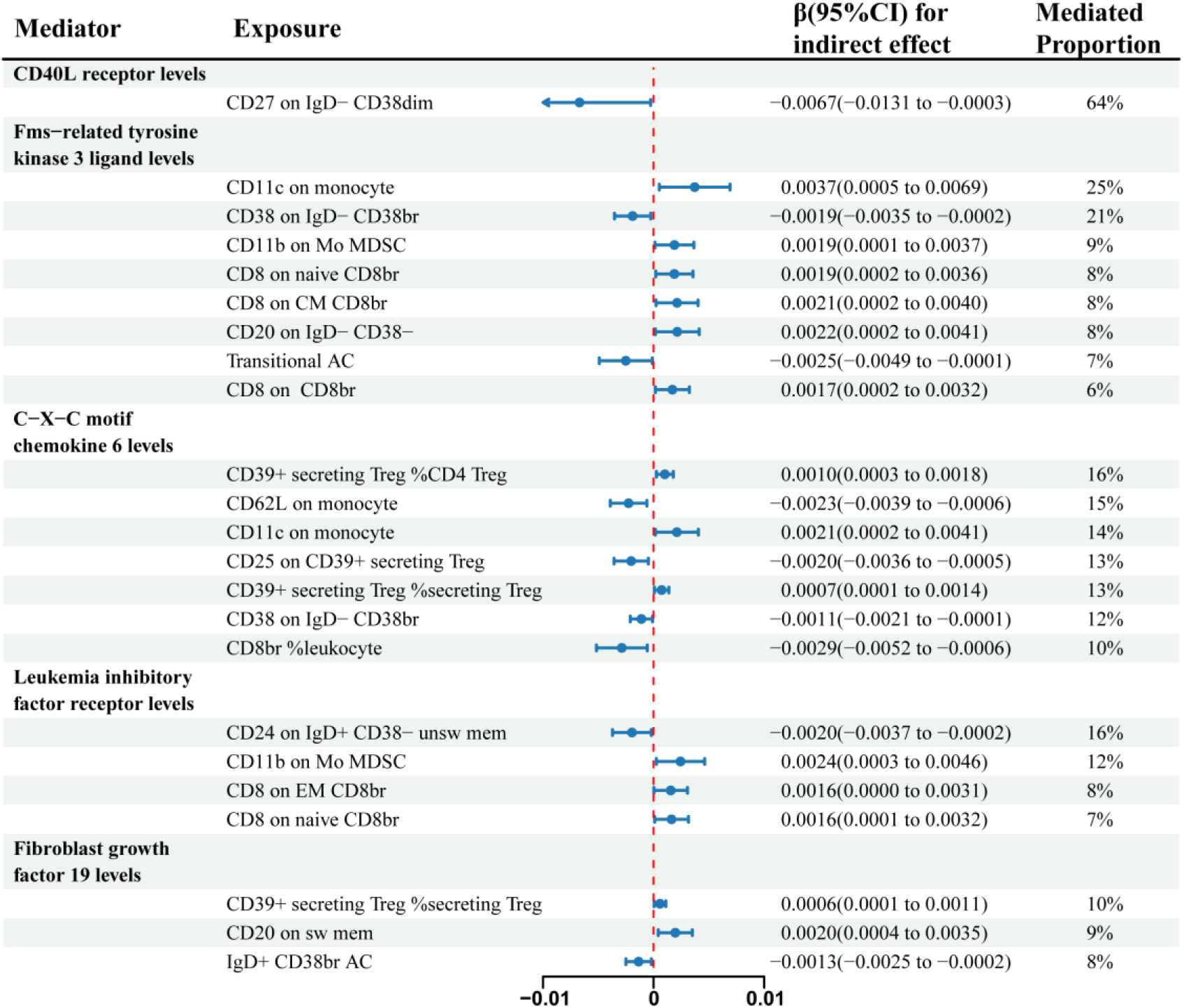
Forest plot for the mediation effect of immune traits on Overall BC via inflammatory protein. **Note:** CI: confidence interval.

**Figure S9.**
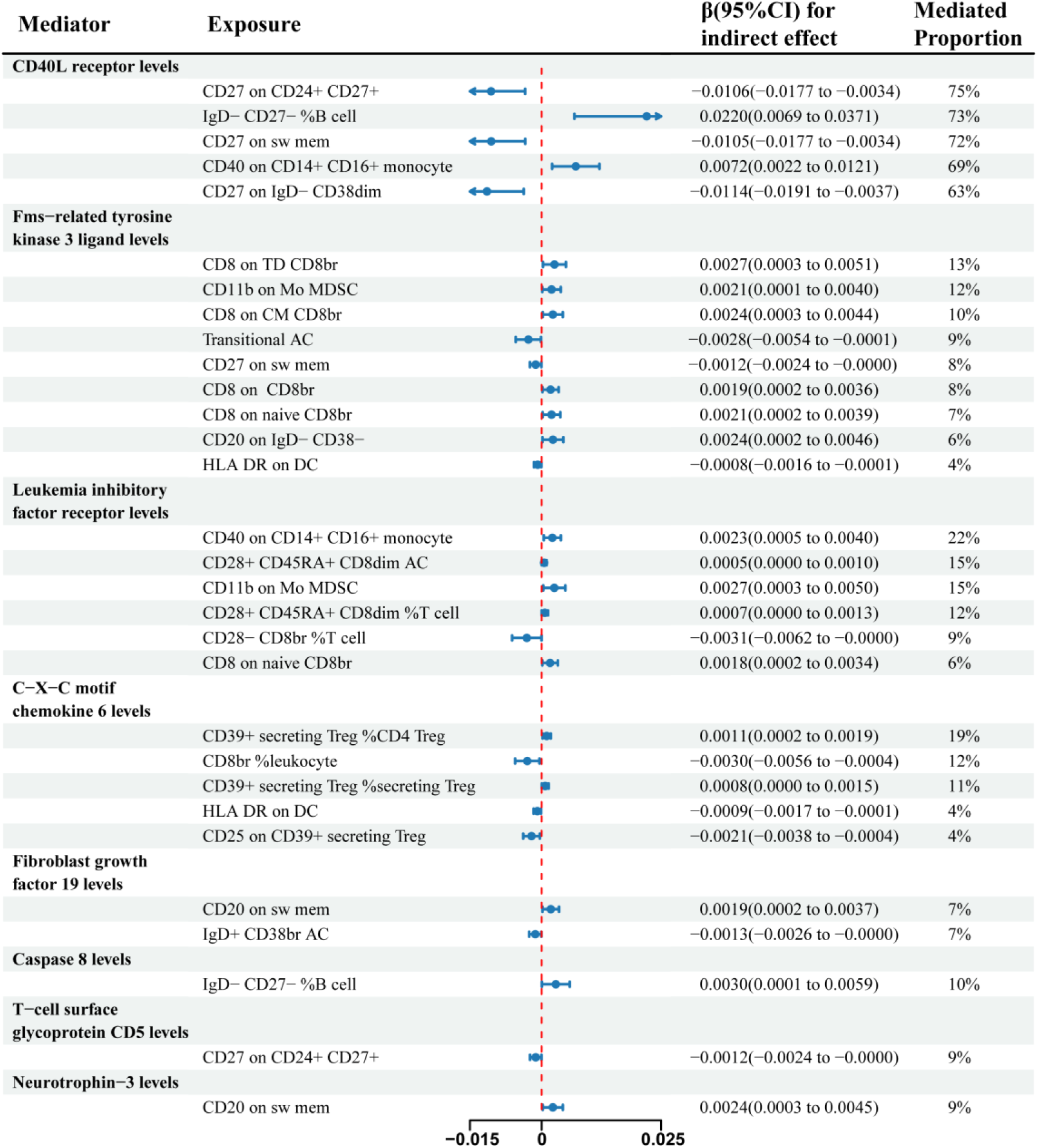
Forest plot for the mediation effect of immune traits on ER+BC via inflammatory protein. **Note:** CI: confidence interval.

**Figure S10.**
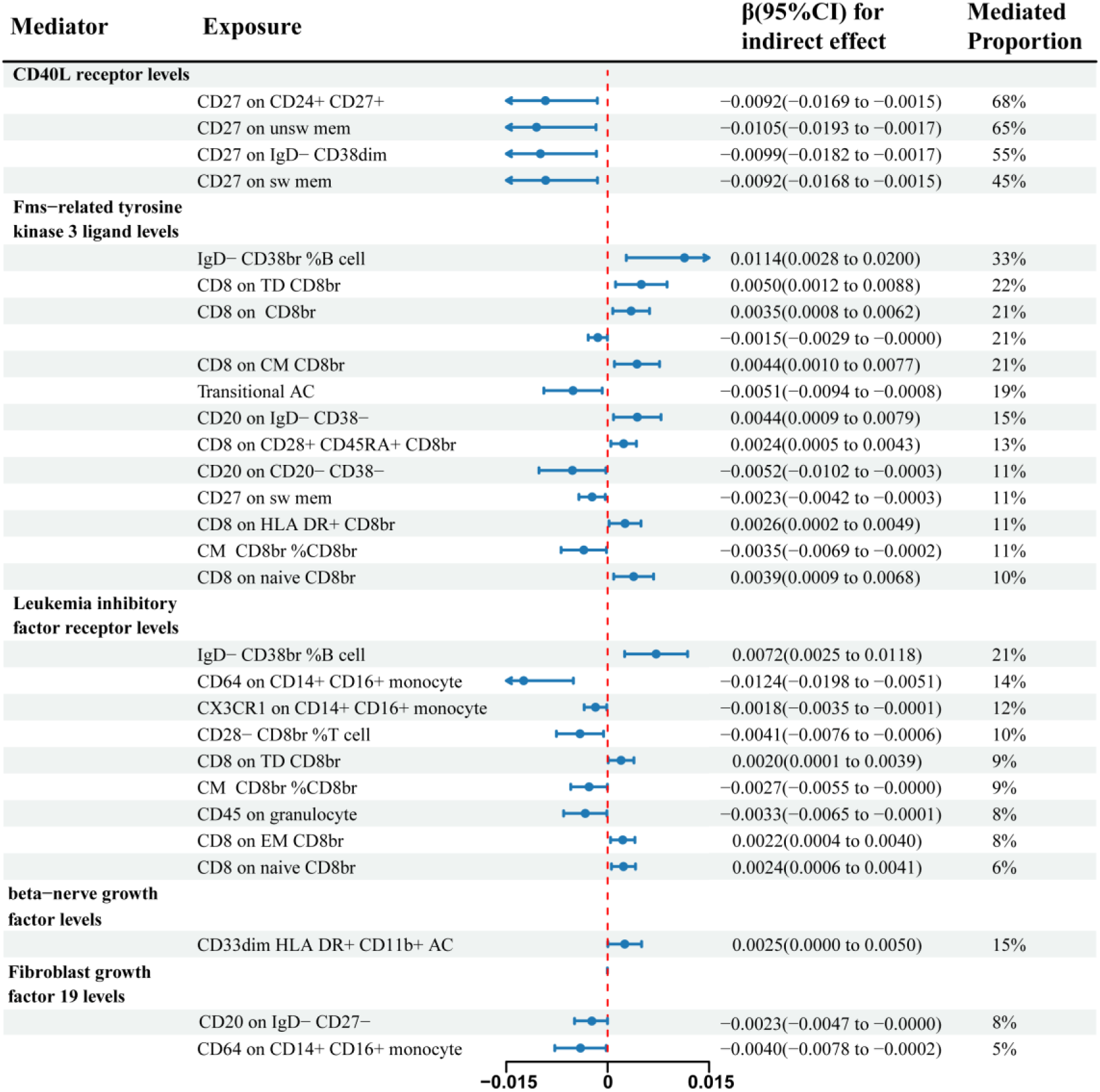
Forest plot for the mediation effect of immune traits on Luminal A like BC via inflammatory protein. **Note:** CI: confidence interval.

**Figure S11.**
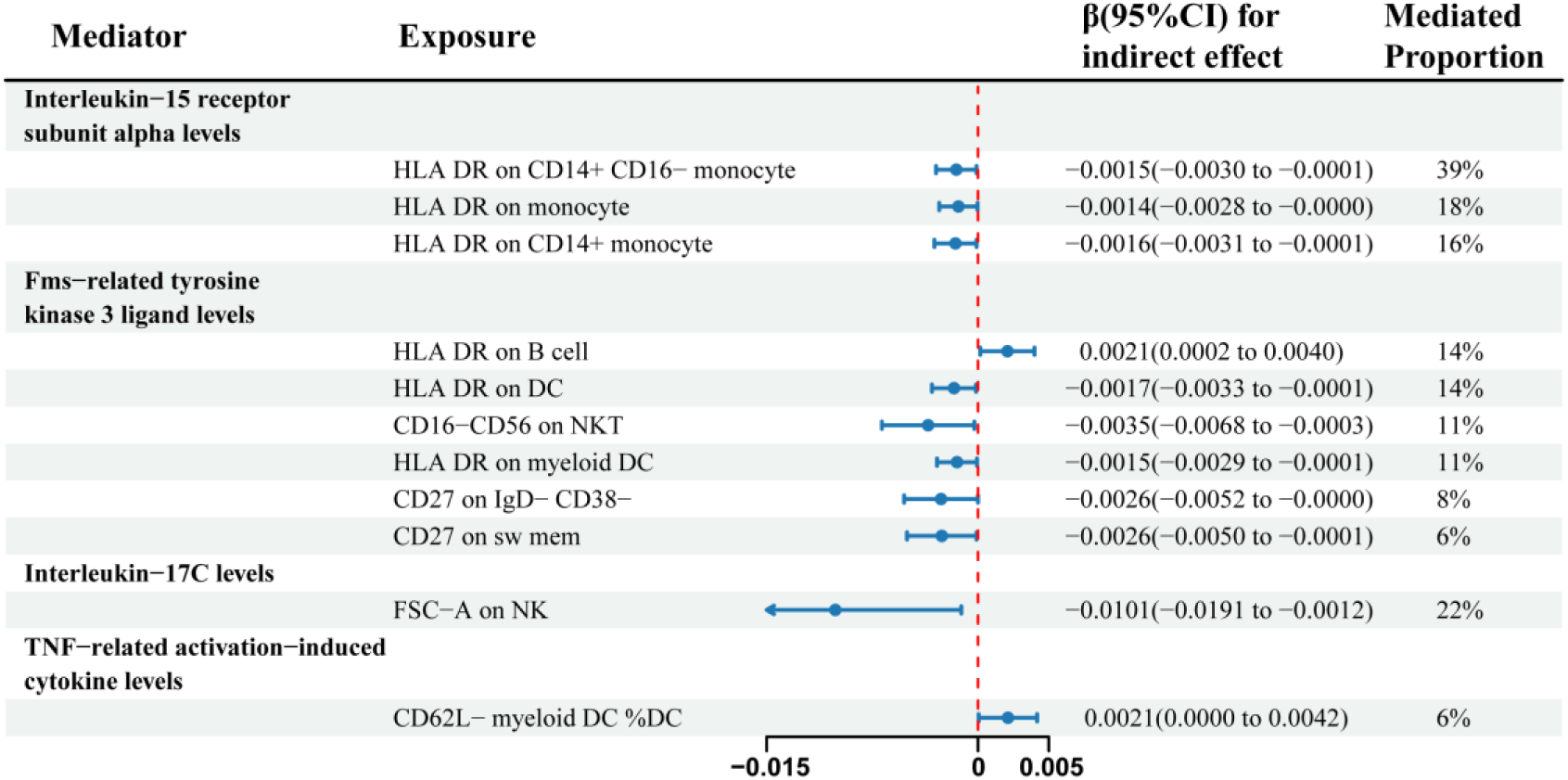
Forest plot for the mediation effect of immune traits on Luminal B/HER2- negative-like BC via inflammatory protein. **Note:** CI: confidence interval.

**Figure S12.**
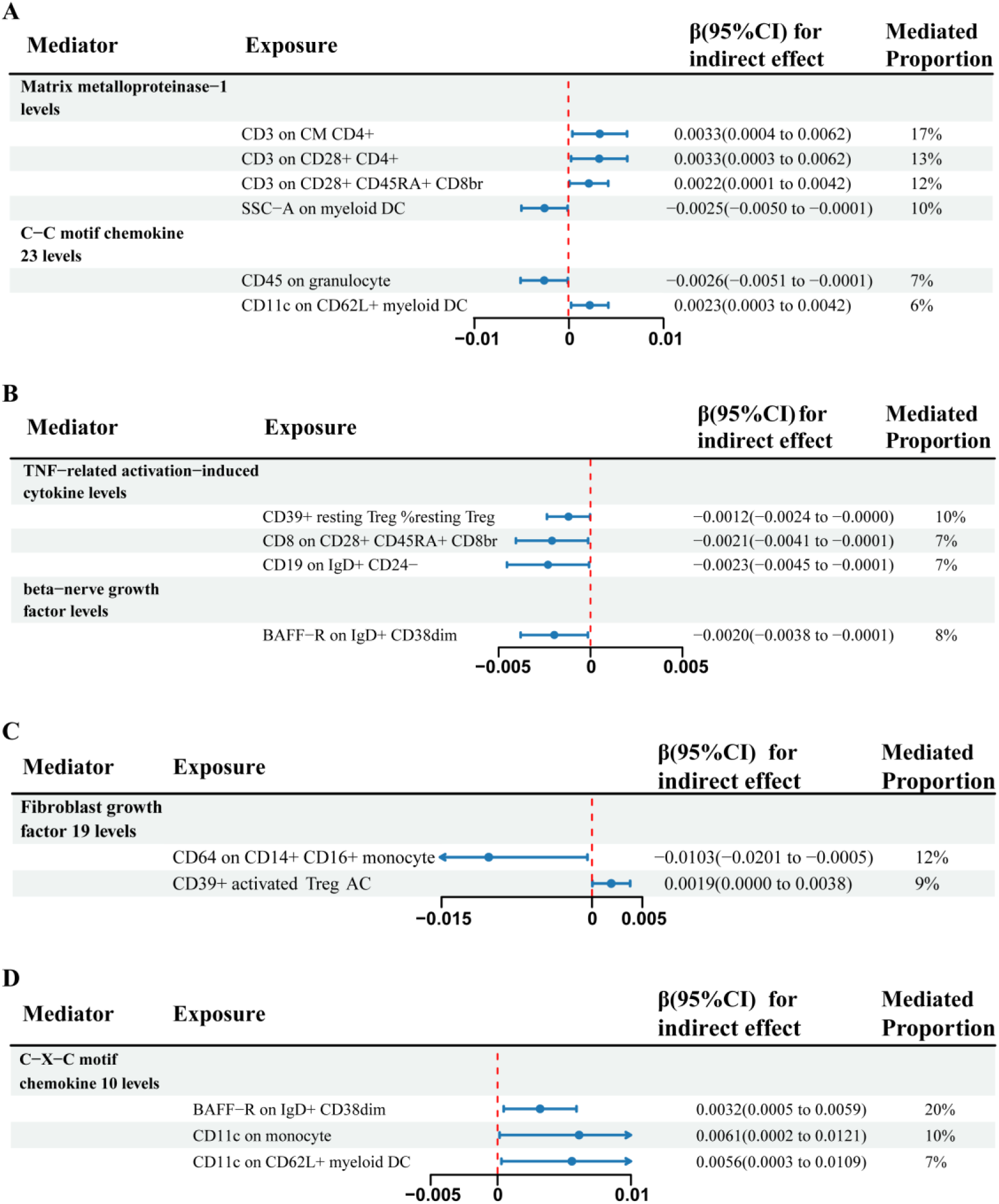
Forest plot for the mediation effect of immune traits on ER-BC, TNBC, Luminal B like BC and HER2-enriched-like BC via inflammatory protein. **Note: (A)** ER-BC; (**B)** TNBC; **(C)** Luminal B like BC; **(D)** HER2-enriched-like BC; CI: confidence interval.

## Notes

### Competing Interest Statement

The authors have declared no competing interest.

